# Autoantibodies linked to autoimmune diseases associate with COVID-19 outcomes

**DOI:** 10.1101/2022.02.17.22271057

**Authors:** Gabriela Crispim Baiocchi, Aristo Vojdani, Avi Z Rosenberg, Elroy Vojdani, Gilad Halpert, Yuri Ostrinski, Israel Zyskind, Igor Salerno Filgueiras, Lena F. Schimke, Alexandre H. C. Marques, Lasse M. Giil, Yael Bublil Lavi, Jonathan I. Silverberg, Jason Zimmerman, Dana Ashley Hill, Amanda Thornton, Myungjin Kim, Roberta De Vito, Dennyson Leandro M. Fonseca, Desireé Rodrigues Plaça, Paula Paccielli Freire, Niels Olsen Saraiva Camara, Vera Lúcia Garcia Calich, Harald Heidecke, Miriam T. Lattin, Hans D. Ochs, Gabriela Riemekasten, Howard Amital, Otavio Cabral-Marques, Yehuda Shoenfeld

**Affiliations:** Department of Immunology, Institute of Biomedical Sciences, University of São Paulo, São Paulo, SP, Brazil; Department of Immunology, Immunosciences Laboratory, Inc., Los Angeles, CA, United States; Cyrex Laboratories, LLC 2602 S. 24th St., Phoenix, AZ 85034 USA; Department of Pathology, Johns Hopkins University, Baltimore, Maryland, USA; Regenera Medical 11860 Wilshire Blvd., Ste. 301, Los Angeles, CA 90025 USA; Ariel University, Israel; Zabludowicz Center for Autoimmune Diseases, Sheba Medical Center, Tel-Hashomer, Israel; Saint Petersburg State University Russia; Department of Pediatrics, NYU Langone Medical Center, New York, NY, USA; Maimonides Medical Center, Brooklyn, NY, USA; Department of Internal Medicine, Haraldsplass Deaconess Hospital, Bergen, Norway; Department of Chemistry Ben Gurion University Beer-Sheva, 84105, Israel; Department of Dermatology, George Washington University School of Medicine and Health Sciences, Washington, DC, USA; ResourcePath, Sterling, VA; Data Science Initiative at Brown University, Providence, RI, USA; Department of Biostatistics and the Data Science Initiative at Brown University, Providence, RI, USA; Department of Clinical and Toxicological Analyses, School of Pharmaceutical Sciences, University of São Paulo, São Paulo, SP, Brazil; CellTrend Gesellschaft mit beschränkter Haftung (GmbH), Luckenwalde, Germany; Department of Biology, Yeshiva University, Manhatten, NY, USA; Department of Pediatrics, University of Washington School of Medicine, and Seattle Children’s Research Institute, Seattle, WA, USA; Department of Rheumatology, University Medical Center Schleswig-Holstein Campus Lübeck, Lübeck, Germany; Department of Medicine B, Sheba Medical Center, Tel Hashomer, Israel; Sackler Faculty of Medicine, Tel-Aviv University, Tel-Aviv, Israel; Network of Immunity in Infection, Malignancy, and Autoimmunity (NIIMA), Universal Scientific Education and Research Network (USERN), São Paulo, SP, Brazil

**Keywords:** Autoantibodies, autoimmune diseases, SARS-CoV-2 infection, COVID-19 outcomes

## Abstract

The SARS-CoV-2 infection is associated with increased levels of autoantibodies targeting immunological proteins such as cytokines and chemokines. Reports further indicate that COVID-19 patients may develop a wide spectrum of autoimmune diseases due to reasons not fully understood. Even so, the landscape of autoantibodies induced by SARS-CoV-2 infection remains uncharted territory. To gain more insight, we carried out a comprehensive assessment of autoantibodies known to be linked to diverse autoimmune diseases observed in COVID-19 patients, in a cohort of 248 individuals, of which171 were COVID-19 patients (74 with mild, 65 moderate, and 32 with severe disease) and 77were healthy controls. Dysregulated autoantibody serum levels, characterized mainly by elevated concentrations, occurred mostly in patients with moderate or severe COVID-19 infection, and was accompanied by a progressive disruption of physiologic IgG and IgA autoantibody signatures. A similar perturbation was found in patients with anosmia. Notably, autoantibody levels often accompanied anti-SARS-CoV-2 antibody concentrations, being both indicated by random forest classification as strong predictors of COVID-19 outcome, together with age. Moreover, higher levels of autoantibodies (mainly IgGs) were seen in the elderly with severe disease compared with young COVID-19 patients with severe disease. These findings suggest that the SARS-CoV-2 infection induces a broader loss of self-tolerance than previously thought, providing new ideas for therapeutic interventions.

## Introduction

Since the first case in December 2019, more than 395 million confirmed cases of the Coronavirus disease 2019 (COVID-19), including 5,7 million deaths, were reported^1^. COVID-19 is caused by the severe acute respiratory syndrome coronavirus 2 (SARS-CoV-2), a single-stranded RNA virus that belongs to the Coronaviridae family^2^. Subsequently, new SARS-CoV-2 variants have kept on emerging, each presenting with selective advantages^3–5^ and threatening the possibility to dominate the COVID-19 pandemic^6^. Concurrently, a massive scientific endeavor to understand COVID-19 pathophysiology has found evidence that patients with severe COVID-19 may present with systemic immune dysregulation of both the innate and adaptive immune responses. This manifests mainly as cytokine storm syndrome^7^, hyperactivation of both myeloid and lymphoid cells^7,8^, characterized by neutrophilia and lymphopenia)^9^.. In addition, recent reports from our group and others^10,11,12^ indicate that patients with COVID-19 have high levels of autoantibody production, suggesting that SARS-CoV-2 can trigger features resembling systemic autoimmune disease^13–17^.

In healthy individuals, autoantibodies are present at physiological levels regulating a myriad of physiological functions, which are essential to maintain body homeostasis^18–21^. However, abnormal levels of autoantibodies often predict preclinical autoimmune diseases^22,23^ as well as promote or exacerbate inflammatory conditions by triggering or exacerbating inflammation, dysregulating biological pathways, and inducing cell lysis^24^. Therefore, autoantibody levels in COVID-19 may have clinical implications. The identified link between SARS-CoV-2 and altered autoantibody profiles suggest that similar to other viruses^25–28^, SARS-CoV-2 infection may result in a life-threatening generalized loss of self-tolerance^13–15,29^. This includes antiphospholipid syndrome^30^, Gillian-Barré syndrome^31^, polyneuritis cranialis^32^, new-onset type 1 diabetes ^33^, autoimmune hemolytic anemia^34^, Graves’ disease^35^, immune thrombocytopenic purpura^36^, arthritis^37^, and systemic lupus erythematosus^16,38^. However, the full spectrum of autoantibodies observed in COVID-19 patients which could indicate a broad loss of self-tolerance, has not been fully characterized. Among the recently reported autoantibodies identified in COVID-19 are those targeting immune related proteins such as type I interferons (IFNs)^11^, the exoproteome (cytokine, chemokine, and their receptors as well as complement factors)^10^, G protein-coupled receptors (GPCRs) and renin-angiotensin system (RAS)-related molecules^12^.

While a select group of autoantibodies observed in classical autoimmune diseases have been associated with COVID-19 severity such as anti-nuclear antibodies (ANAs^39^), ribosomal P proteins, chromatin proteins, and thyroid antigens^40^ although comprehensive studies broadly characterizing autoantibodies induced by SARS-CoV-2 are lacking. Furthermore, high levels of antiphospholipid autoantibodies were linked to severe COVID-19 by inducing neutrophil extracellular traps (NETs) and venous thrombosis^16,41–44^. Based on these observations, autoantibodies associated with diverse autoimmune diseases and the loss of self-tolerance might be associated with the severity of COVID-19 as previously indicated^14,16,17,40,45,46^ and by extension suggest that loss of self-tolerance is a feature of severe COVID-19. To better characterize the immunopathology of COVID-19 it is imperative to determine the landscape of autoantibodies associated with the severity of SARS-CoV-2 infection, which will lead to improve therapeutic interventions. To achieve this goal, we have characterized the spectrum of autoantibodies linked to autoimmune diseases in patients with different COVID-19 clinical outcomes, using a systems immunology approach.

## METHODS

### Study Participants

A total of 248 adults belonging to Jewish communities from 5 states of the United States of America were enrolled in this study. Of these, 171 patients had COVID-19 symptoms (**Supplementary Table 1**) and a positive SARS-CoV-2 test by nasopharyngeal swab and PCR analysis (polymerase chain reaction) before receiving any SARS-CoV-2 vaccine. These patients participated in an online survey developed to determine the most common symptoms and outcomes of SARS-CoV-2 infection^47,48^. The SARS-CoV-2 positive cohort was classified as mild COVID-19 (n=74; fever duration ≤ 1 day; peak temperature of 37.8 C), moderate COVID-19 (n=63; fever duration ≥ 7 days; peak fever of ≥ 38.8 C), and severe COVID-19 groups (n=32; severe symptoms and requiring supplemental oxygen therapy) according to the World Health Organization (WHO) severity classification^49^. Details of the survey study, patient demographics, and symptoms were previously described^47,48^ and can be found in **Supplementary Table 1**. We also included 77 randomly selected age- and sex-matched healthy controls from the same Jewish population who were SARS-CoV-2 negative and did not present any COVID-19 symptoms. All healthy controls and all patients provided informed written consent to participate in the study, which was performed in accordance with the Declaration of Helsinki. The study was approved by the IntegReview institutional review board (Coronavirus Antibody Prevalence Study, CAPS-613). In addition, this study followed the reporting guidelines of Strengthening the Reporting of Observational Studies in Epidemiology ([STROBE](https://www.equator-network.org/reporting-guidelines/strobe/)).

### Measurements of anti-SARS-CoV-2 antibody and autoantibodies linked to autoimmune diseases

All collected sera were tested for IgG anti-SARS-CoV-2 antibody using the ZEUS SARS-CoV-2 ELISA Test System according to the manufacturer’s instructions (ZEUS Scientific, New Jersey, USA). This is an ELISA test that measures IgG antibodies to spike and nucleoprotein combined. Serum IgG autoantibodies against nuclear antigen (ANA), extractable nuclear antigen (ENA), double-stranded DNA (dsDNA), actin, mitochondrial M2, and rheumatoid factor (RF) were measured using commercial ELISA kits obtained from INOVA Diagnostics (San Diego, CA, USA). In addition, IgG and IgA antibodies against 52 different autoantigens^50–58^ were measured by in-house ELISA procedure at Immunosciences Lab., Inc. (Los Angeles, CA USA) in blinded fashion. See **Supplementary Table 2** for the autoantigens tested.. For the measurement of these antibodies, one hundred mL of each autoantigen at the optimal concentration, prepared in 0.01 M PBS pH 7.4 were added to a series of microtiter plates. A set of plate wells were also coated with 2% bovine serum albumin (BSA) or human serum albumin (HSA) and used as controls. The ELISA plates were incubated overnight at 4°C and then were washed five times with 250 ml of 0.01 M PBS containing 0.05% Tween 20 pH 7.4. The non-specific binding of immunoglobins was prevented by the addition of 200 mL of 2% BSA in PBS, which was then incubated overnight at 4°C. Plates were washed and the serum samples from healthy controls and the SARS-CoV-2 patients were diluted at 1:50 for the determination of IgA antibody, and 1:100 for the determination of IgG antibody in serum diluent buffer or 1% BSA in PBS containing 0.05% Tween 20 was added to the wells of ELISA plates, which were then incubated for one hour at room temperature. Plates were washed, followed by the addition of alkaline phosphatase goat anti-human IgG or IgA F(ab,)2 fragments (KPI, Gaithesburg, MD, USA) at an optimal dilution of 1:200 for IgA and 1:600 for IgG in 1% BSA-PBS. The plates were then incubated for another hour at room temperature. After five washings with PBS-Tween buffer, the enzyme reaction was started by adding 100 mL of para-nitrophenylphosphate in 0.1 mL diethanolamine buffer 1 mg/mL containing 1 mM MgCl_2_ and sodium azide pH 9.8. Forty-five minutes later, the reaction was stopped with 50 mL of 1 N NaOH. The optical density (OD) was read at 405 nm with a microtiter plate reader. The ODs of the control wells containing only HSA or BSA which were less than 0.15 were subtracted from all other wells to exclude non-specific binding. The ELISA index for each antibody was calculated and the background of the ELISA assay was not more than 0.150 O.D.

### Multistudy factor analysis

Multistudy factor analysis (MSFA) was performed using the R package called MSFA^59^, which uses either a fast expectation conditional-maximization algorithm and a Gibbs Sampling algorithm^60^ for the parameter estimate. Here, we adopt the Gibbs Sampling algorithm to estimate the parameter in a Bayesian framework^60^. MSFA is an unsupervised and inferential analysis used to identify common and study-specific factors shared by the control and COVID-19 groups as previously described^61^.

### Differences in autoantibody levels

Box plots showing differences in autoantibodies from COVID-19 patient groups (mild, moderate and severe) and healthy controls were generated using the R version 4.0.5 (The R Project for Statistical Computing. https://www.r-project.org/), R studio Version 1.4.1106 (R-Studio. https://www.rstudio.com/), and the R packages ggpubr, lemon, and ggplot2. Statistical differences in autoantibody levels were assessed using a two-sided Wilcoxon rank-sum^10^.

### Principal component analysis (PCA)

PCA with spectral decomposition^62,63^ was carried out to measure the stratification power of the autoantibodies in distinguishing COVID-19 (mild, moderate, and severe patients) and healthy controls. PCA was performed using the R functions prcomp and princomp, through the factoextra package (PCA in R: prcomp vs princomp)^64^.

### Machine learning model and autoantibody ranking

We employed random forest model to construct a classifier for discriminating controls and mild, moderate, and severe COVID-19 patients. The aim of this approach was to identify the most significant predictors for severe COVID-19. We trained the random forest model using the functionalities of the R package randomForest (version 4.6.14)^65^. Five thousand trees were used, and 3 variables were resampled. A follow-up analysis was conducted with the Gini decrease, the number of nodes and mean minimum depth as criteria to determine variable importance. The adequacy of the random forest model as a classifier was assessed through the out-of-bags error rate and the ROC curve. For cross-validation, we split the dataset into training and testing sets, using 75% of the observations for training and 25% for testing.

### Autoantibody correlation signatures

Bivariate correlation analyses of autoantibodies for each group (controls and mild, moderate and severe COVID-19 patients) were performed using the corrgram, psych and inlmisc R packages. In addition, circle plots of autoantibody correlation were built using the R packages qgraph, ggplot2, psych, inlmisc to visualize the patterns of Spearman’s rank correlation coefficients between autoantibodies.

## RESULTS

### Progressive increase of autoantibodies in COVID-19 patients according to disease severity

Here we investigated the serum levels of IgG and IgA autoantibodies targeting 58 and 52 self-antigens, respectively, that are linked to a variety of autoimmune diseases (**Fig. 1)**. See **Supplementary Table 1** for autoantibody levels, **Supplementary Table 2** for their abbreviations and targets, and **Supplementary Table 3** for autoantibody and disease association. Multi-study factor analysis (MSFA) (**Fig. 2 and Supplementary Table 6**) revealed a progressive dysregulation in the number of latent factors from mild COVID-19 patients to those with moderate and severe disease. The two latter groups presented fewer latent factors when compared with control and mild COVID-19 groups, indicating by this statistical inference approach that the perturbation of normal autoantibody levels mostly occurs in patients with moderated and severe COVID-19.

**Figure 1.**
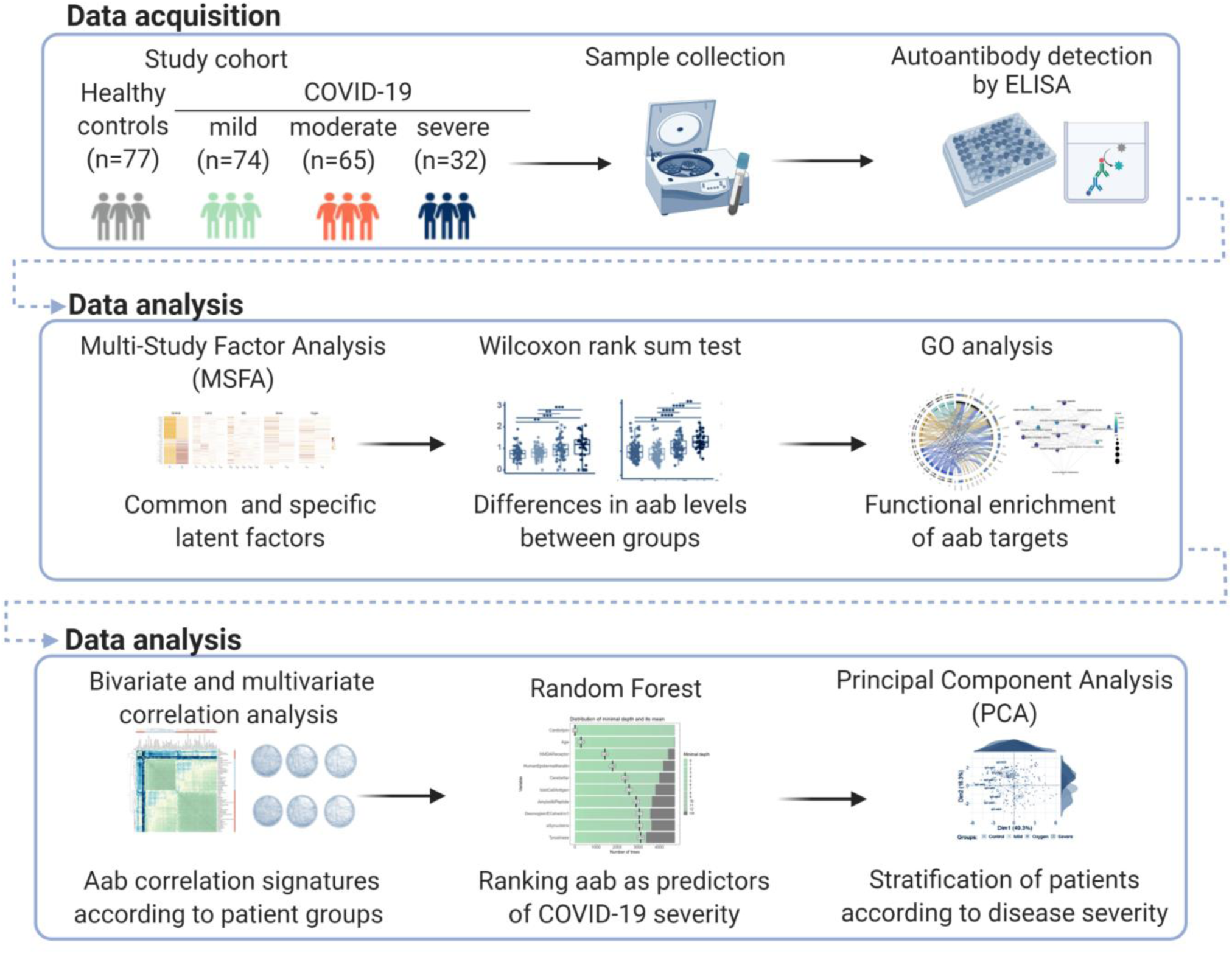
Study workflow. **a)** Schematic overview of study cohorts, statistics and bioinformatics analyses carried out to characterize the spectrum of autoantibodies linked to diverse autoimmune diseases that associate with COVID-19 outcomes.

**Figure 2.**
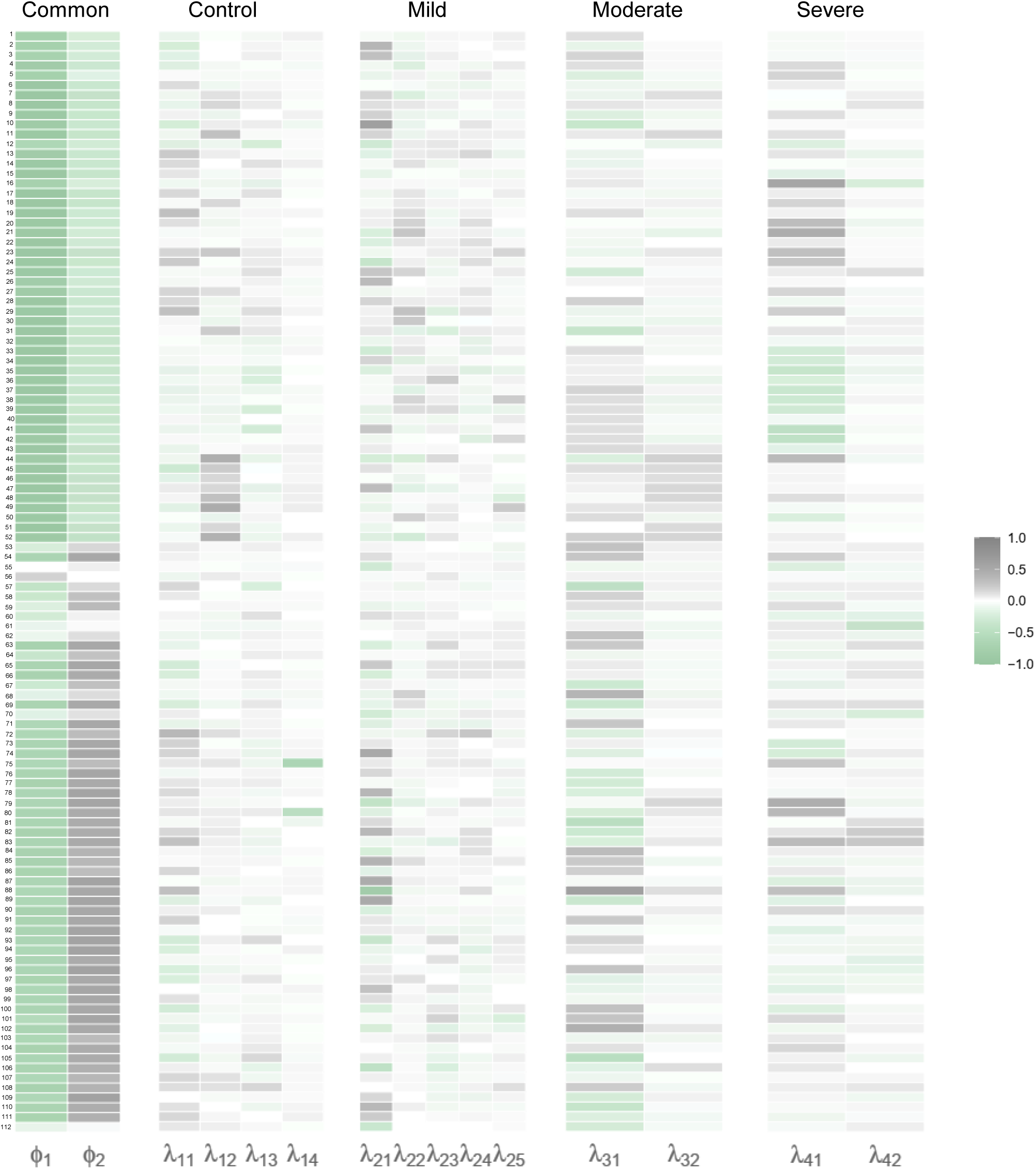
Perturbation of normal autoantibody settings in COVID-19. Multi-study factor analysis (MSFA) of all IgA and IgG autoantibodies as well as anti-SARS-CoV-2 IgG antibodies that were analyzed. The heatmaps show estimated factor loadings of common (Φ) and specific (λ) latent factors of each group. Positive and negative loadings range from − 1 (green) to 1 (grey), respectively. Autoantibody number shown according to their names in **Supplementary Table 1**. See **Supplementary Table 6** for the loadings.

We found significantly elevated levels of IgG (**Supplementary Fig. 1)** and IgA (**Supplementary Fig. 2**) autoantibodies targeting a total of 42 and 25 antigens, respectively, when comparing COVID-19 groups (mild, moderate, and severe) versus healthy controls (**Fig. 3a**). In general, while patients with mild COVID-19 exhibited only slight differences in the serum levels of autoantibodies when compared to healthy controls, patients with moderate COVID-19 exhibited more evident differences, with the highest levels of autoantibodies noted in the severe group. In agreement with the results obtained by MSFA, we observed a progressive increase of the autoantibodies targeting autoantigens such as those associated with Alzheimer’s disease (rabaptin-5, tau protein), antiphospholipid syndrome (cardiolipin), multiple sclerosis and other neuroimmune disorders (myelin basic protein), celiac disease (transglutaminases, zonulin), neuropsychiatric and neurodegenerative disorders (NMDAR), gut motility disorders (enteric nerve), liver autoimmunity (liver microsomal antigen), immune thrombocytopenia (platelet glycoprotein),myasthenia gravis (acetylcholine receptor, somatotropin), neurodegenerative diseases (neurofilament proteins), Parkinson’s disease (α-synuclein), pemphigus vulgaris or foliaceus (Desmoglein-E-Cadherin 1), neurological disorders (S100B), rheumatic heart disease (α-myosin), rheumatoid arthritis (α-enolase, fibulin), systemic lupus erythematosus (cardiolipin, dsDNA, rabaptin-5), type 1 diabetes (islet cell antigen), immune thrombotic thrombocytopenia (heparin), and vitiligo (tyrosinase) (**Supplementary Table 3, Supplementary Fig. 1-2 and Fig. 3a, 3d-e)**. In addition, we also identified reduced levels of IgG autoantibodies such as those against a-myosin, rabaptin-5, S100B, and cerebellar as well as IgA autoantibodies targeting the NMDA R and cerebellar. Thus, indicating a broad breakdown of physiological autoantibody levels/self-tolerance in patients with COVID-19 that paralleled disease severity. Of note, the progressive perturbation of autoantibody levels was accompanied by increased serum concentration of anti-SARS-CoV-2 antibodies (**Fig. 3e**).

**Figure 3.**
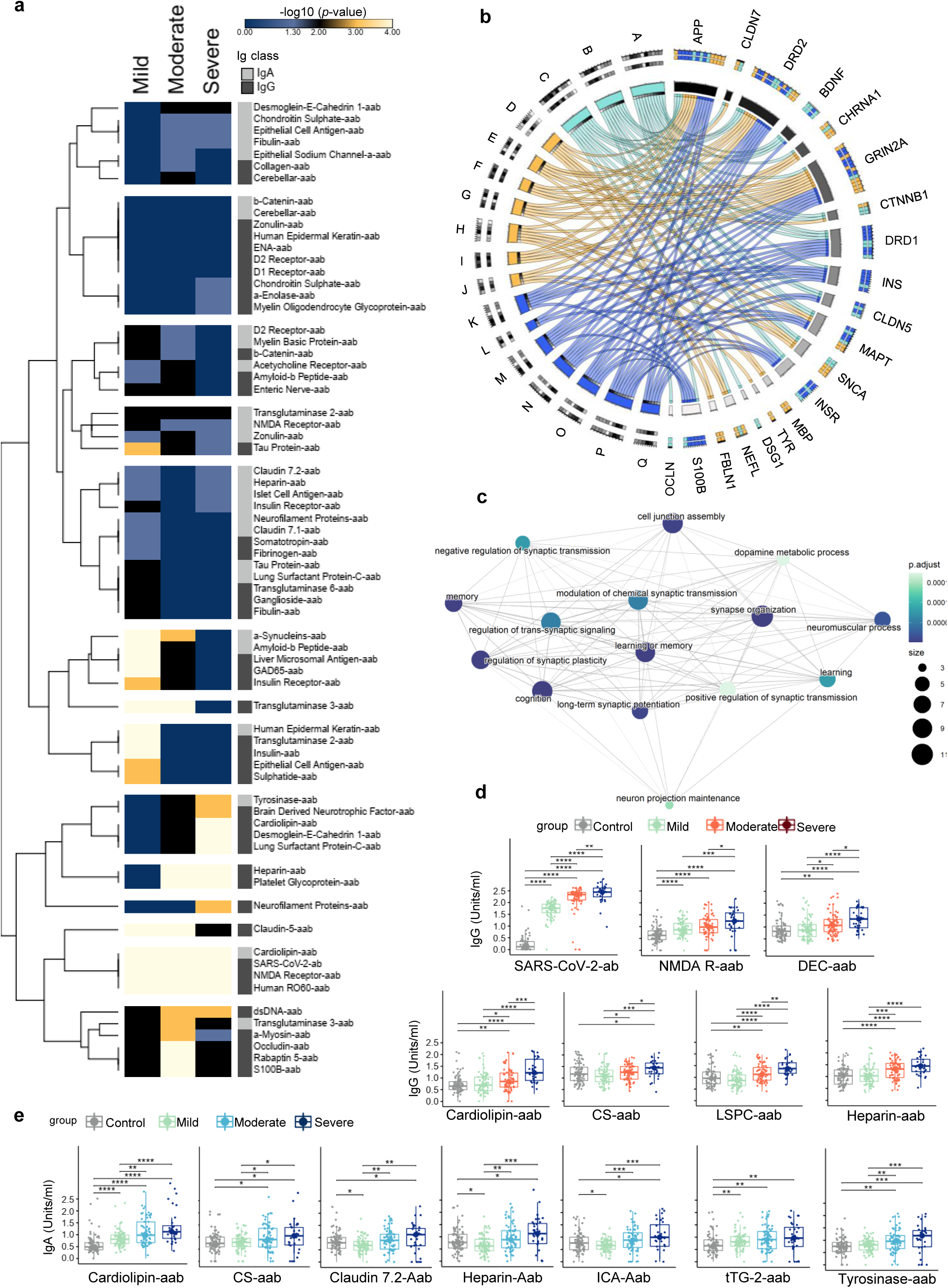
Autoantibodies and functional association of their targets. **a)** Heatmap showing the hierarchical cluster of IgG and IgA autoantibodies significantly different when comparing each COVID-19 cohort (mild, moderate, and severe groups) with healthy controls. The bar, ranging from blue to black and light yellow, shows the range of significance from -log10 (*p-* value) = 0 to 4. Rows were clustered using Euclidean distance between -log10 (*p*-values). Ig, immunoglobulin. **b**) The barplots exhibt the number of IgG and IgA autoantibodies that were evaluated and found as significant or not. **c**) Network of biological processes (BP) enriched (adjusted p-value <0.05) by the autoantibody targets. The edges connect related BPs. The circles’ colors and size correspond to the adjusted *p*-value and number of autoantibody targets enriching BPs, respectively. **d**) Circos plot illustrates the top 10 BPs enriched by both IgA and IgG autoantibody targets. **Suplementary Table 4 and Supplementary Fig. 3** show all gene ontology (GO) categories (BPs and signaling pathways) enriched by the autoantibody targets. For this analysis, we selected autoantibody targets from significantly different autoantibodies among the groups. BPs are denoted by letters A-Q (A: synapse organization; B: learning or memory; C: cell junction assembly; D: positive regulation of peptidase activity; E: response to alcohol; F: excitatory postsynaptic potential; G: neuromuscular process; H: positive regulation of proteolysis; I: chemical synaptic transmission, postsynaptic; J: phenol-containing compound metabolic process; K: long-term synaptic potentiation; L: neuron projection maintenance; M: positive regulation of synaptic transmission; N: regulation of synaptic plasticity; O: memory; P: cognition; Q: learning). The blue, yellow, and green colors correspond to IgA, IgG, and shared terms, respectively. Box plots exemplifying **d)** IgG and **e**) IgA autoantibodies significantly (different between healthy controls and COVID-19 groups. See **Supplementary Table 2** for full names and abbreviations of autoantibodies and their targets. All autoantibodies with significant differences between at least two groups are shown in **Supplementary Figures 1 and 2**. Each box plot shows the median with first and third interquartile range (IQR), whiskers representing minimum and maximum values within IQR, and individual data points. Significance was determined using two-sided Wilcoxon rank-sum tests and is indicated by asterisks (* p ≤ 0.05, ** p ≤ 0.01, *** p ≤ 0.001, and **** p ≤ 0.0001). Control (*n* = 77); mild (*n* = 74), moderate (*n* = 65), and severe (*n* = 32) COVID-19.

**Fig. 3b and Fig. 3c** display the functional associations of the autoantibody targets, which are represented by functional enrichment analysis (gene ontology or GO). These results indicate that, among others, COVID-19 patients have dysregulated levels of autoantibodies affecting in neurological functions such as memory, learning, and cognition^66–68^ (**Supplementary Table 4 and Supplementary Fig. 3**). Taken together, our data indicate that autoantibodies may also be involved with altered learning, memory and neuroplasticity^68^ recently reported during SARS-CoV-2 infection, which promotes a systemic inflammation milieu that might be neurotoxic.

### Autoantibody generation correlates with severe SARS-CoV-2 infection and anosmia

Based on the concept that signatures of autoantibody correlations are associated with physiological and pathological immune homeostasis^10,12,40^, we investigated the correlation between signatures of the IgA and IgG autoantibodies and disease severity. Bivariate correlation analysis revealed (**Supplementary Table 5 and Fig. 4a**) that patients with mild COVID-19 exhibited few differences in the autoantibody correlation signatures when compared to healthy controls. In contrast, patients with moderate COVID-19 started to exhibit new positive and negative correlations between autoantibodies while the severe group displayed the most disparate topological correlation pattern, increasingly with preferentially positive correlations between autoantibodies.

**Figure 4.**
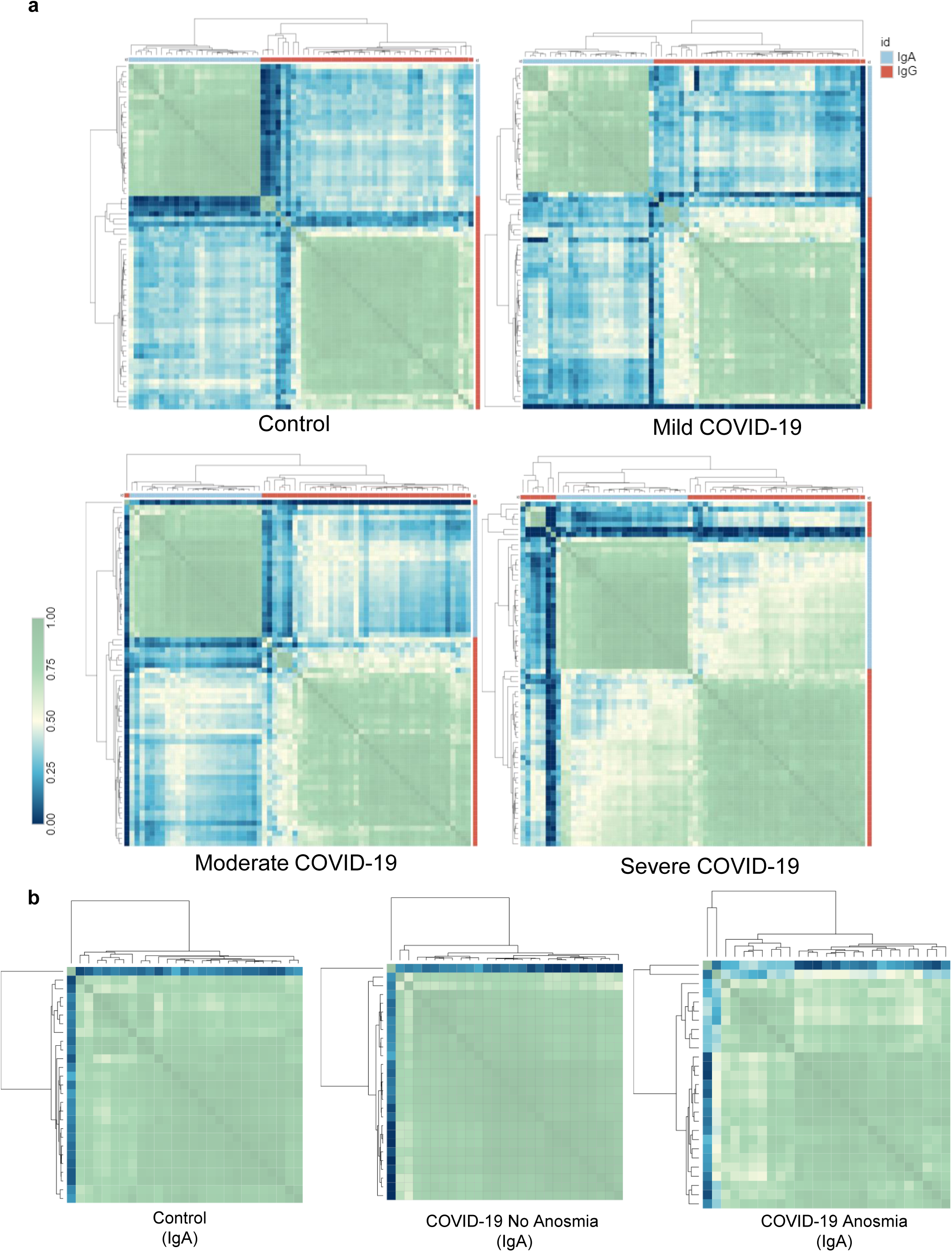
Hierarchical clustering of the autoantibody correlation signatures according to disease severity and anosmia. Heatmap of significant different aab vs. aab correlations: **a)** IgG and IgA autoantibodies in healthy control and COVID-19 groups and **b)** IgA autoantibodies of COVID-19 patients with or without anosmia versus healthy controls. See **Supplementary Fig. 4a-4d** for correlograms with the autoantibody names; **Supplementary Fig. 4e** for IgG autoantibody correlations in patients with anosmia versus no anosmia; and **Supplementary Fig. 5 a-b** for IgG and IgA autoantibodiy correlations for healthy controls and COVID-19 groups. The color scale bar ranging from 0 to 1 represents Spearman’s rank correlation coefficient. Rows and columns were clustered based on Euclidean distance. See **Supplementary Table 5** for correlations matrices with the correlation coefficient values. Control (*n* = 77); mild (*n* = 74), moderate (*n* = 65), and severe (*n* = 32) COVID-19. No anosmia (*n* = 69) and anosmia (*n* = 101).

We then assessed whether anosmia (loss of smell), which was one of the most consistent symptoms associated with SARS-CoV-2 infections^69^, also correlated with autoantibody signatures in patients with mild or moderate COVID-19. Due to the extreme clinical condition requiring oxygen replacement therapy that precludes the characterization of anosmia, patients with severe COVID-19 were excluded from this analysis. We observed that the interconnections among the autoantibodies are reduced in both mild and moderate COVID-19 with anosmia when compared with those without (**Supplementary Fig. 4e and Fig. 3b)**. Taken together, this suggests the presence of multilayered factors that influence the autoantibody signatures, including both disease severity and anosmia that affect relationships among autoantibodies differently.

### Ranking autoantibodies as predictors of COVID-19 severity

To investigate the classification power of autoantibodies, we used random forest modeling, which predicts outcomes based on decision trees^70^, to rank the top 10 autoantibodypredictors of COVID-19 outcomes. Random forest classification of COVID-19 patients presented a stable curve showing the highest error rate (out-of-bag OOB) for the severe COVID-19 group based on both IgA or IgG autoantibodies (**Supplementary Fig. 6a, 6d, 6g, and 6h**). The confusion matrix showed that the random forest model was unable to distinguish moderate from severe COVID-19 groups (**Supplementary Fig. 6b and 6e**). These results indicate that moderate and severe COVID-19 patients may present a similar autoantibody signature. Therefore, we lumped moderate and severe COVID-19 patients into the same group. Using this approach, the out-of-bag (OOB) estimate of the error rate for all groups changed from 37% to 22% for IgA (**Fig. 5d**) and from 25% to 12% for IgG autoantibodies (**Fig. 5a)**. This way, the model was considered adequate to classify the COVID-19 patients by severity according to IgA and IgG autoantibodies, showing areas under the ROC curve of 0.91%, 0.85%, and 0.90% for the healthy control, moderate, and severe COVID-19 groups for IgA (**Supplementary Fig. 6j**) and 0.98%. 0.96% and 0.99% for IgG (**Supplementary Fig. 6i**). In this context, autoantibodies tracked with anti-SARS-CoV-2 antibodies and age as predictive factors for COVID-19 outcome. Among them are IgG autoantibodies targeting the NMDA R, insulin, PG, LSPC, human RO60, claudin, heparin, and S100B, as well as IgA autoantibodies targeting cardiolipin, the NMDA R, HEK, cerebellar, ICA, AbP, DEC, α-synucleins, and tyrosinase. Thus, IgA and IgG autoantibodies stratify COVID-19 according to disease severity.

**Figure 5.**
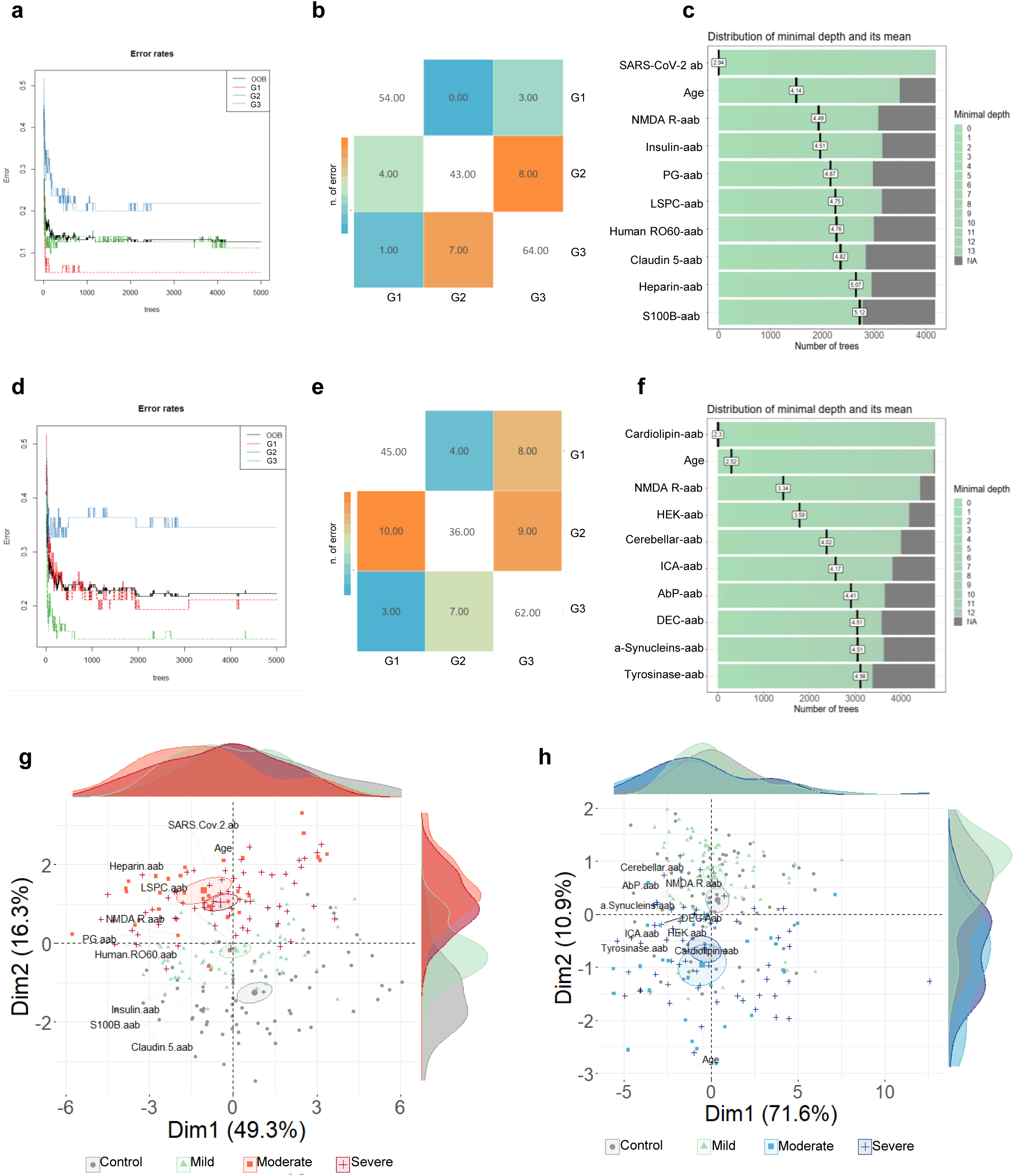
Ranking autoantibodies as predictors of COVID-19 severity. Stable curve showing number of trees and error rates for **a**) IgG and **d**) IgA autoantibodies with out-of-bag (OOB) estimate of 12,5% and 22.28% error rate, respectively. Heatmap presents the confusion matrix of **b**) IgG and **e**) IgA autoantibodies, the number of individuals classification by group (healthy controls = G1; mild = G2; moderate+severe = G3). The color scale ranging from blue to orange corresponds to the number of errors or hits by groups denoted inside the colored or white squares, respectively. Control (*n* = 77); mild (*n* = 74), moderate+severe (*n* = 97) COVID-19. Bar plots of the top 10 strongest **c**) IgG (also anti-SARS-CoV-2 IgG antibodies and age) and **f**) IgA autoantibodies predictors of COVID-19 patient classification according to disease severity. The variables are shown according to minimal depth and number of trees. The color scale bar ranging from 0 to 13 represents the minimal and maximum minimal depth. The small dark vertical bars represent the mean of minimal depth for each variable. Principal component analysis (PCA) with spectral decomposition shows that **g**) IgG and **h**) IgA autoantibodies stratify COVID-19 patients according to disease severity, revealing the overlap between the moderate and severe cohorts. The PCA was carried out based on the top 10 most important variables as listed in **c** and **f**, respectively. Control (*n* = 77); mild (*n* = 74), moderate (*n* = 65), and severe (*n* = 32) COVID-19. See **Supplementary Table 2** for full names and abbreviations of autoantibodies and their targets.

To better understand the stratification power of the autoantibody predictors of COVID-19 outcomes, we performed principal component analysis (PCA) with a spectral decomposition approach^40,41^ to analyze whether the top 10 ranked variables (IgA and IgG autoantibodies and age) as predictors of COVID-19 severity have stratification power. This approach indicated, in accordance with the random forest modeling, that the ranked autoantibodies stratify COVID-19 patients according to disease severity. (**Fig. 5g and 5h**). While healthy controls and patients with mild COVID-19 presented with similar autoantibody patterns, moderate and severe COVID-19 groups clustered closely. Together, these results indicate that the immune responses against SARS-CoV-2 not only are associated with an increase of serum autoantibody levels linked to autoimmune diseases, but also have predictive stratification values for COVID-19 patients.

### SARS-CoV-2 infection dysregulates autoantibody levels in an age-dependent manner

Considering age as a major predictor of severe outcomes in COVID-19, which was also identified by our data (Random forest)^71^, we investigated the effect of age on f the top IgG and IgA autoantibody predictors of COVID-19 severity. We classified control and disease groups by age (young < 60 and elderly ≥ 60) and found that both moderately and severely affected elderly COVID-19 patients show an overall tendency to have elevated autoantibody levels when compared with younger patients (**Fig. 6a**). However, only elderly patients with severe COVID-19 presented with significant differences in most of the top 10 IgG (**Fig. 6b**) and IgA (**Fig. 6c**) autoantibodies when compared with young patients with severe disease. Of note, except for autoantibodies targeting α-enolase, cardiolipin, CS, DEC, dsDNA, ENA, elderly patients with severe COVID-19 had significant differences in all other IgG autoantibodies (**Supplementary Fig. 7a**). In turn, IgA autoantibodies against 10 antigens displayed significant differences comparing young COVID-19 patients to severe COVID-19 patients (**Supplementary Fig. 7b**). No age-dependent differences were observed regarding the levels of anti-SARS-CoV-2 antibodies.

**Figure 6.**
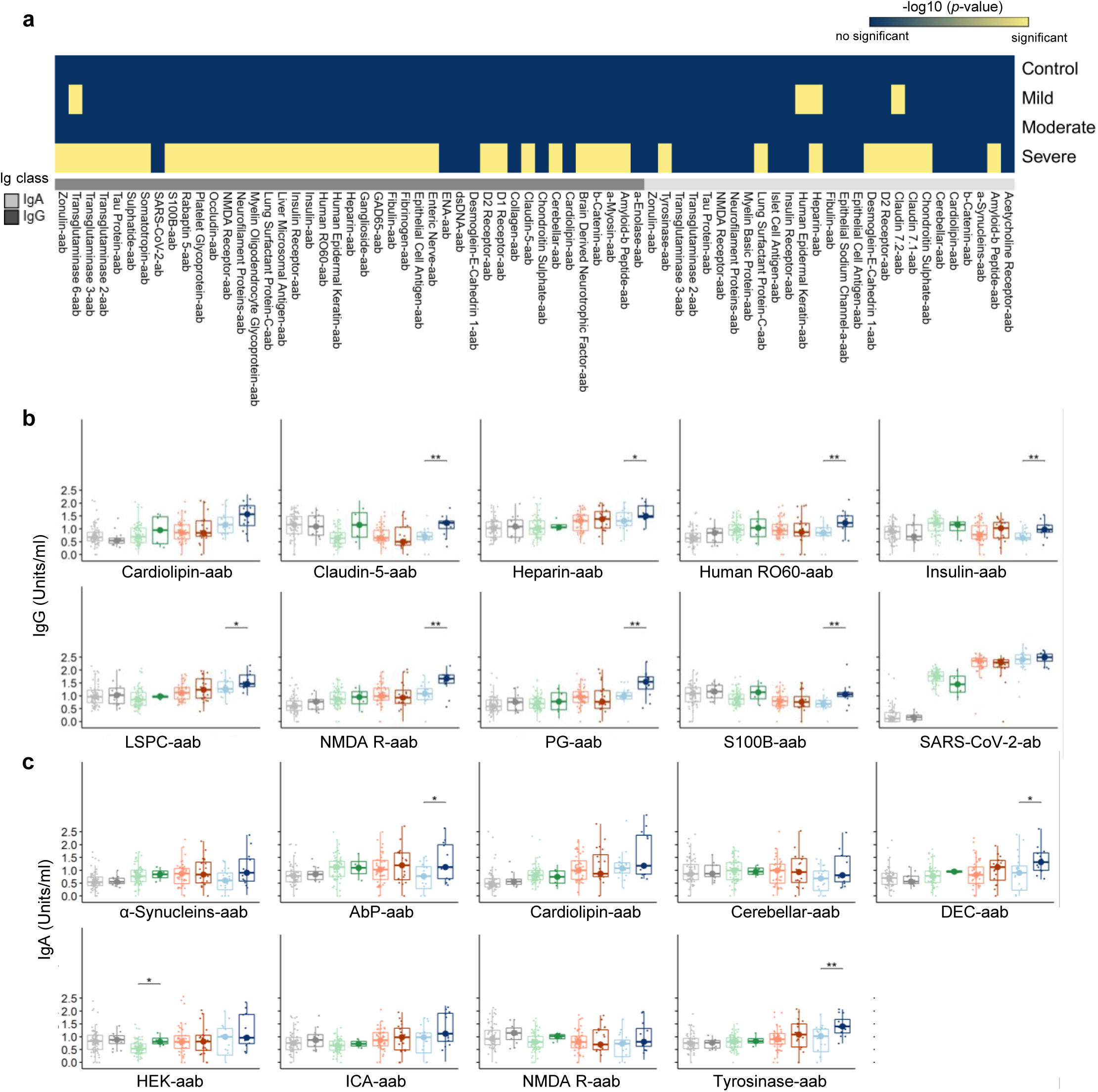
Age-dependent elevation of autoantibody levels in patients with severe SARS-CoV-2 infection. **a)** Heatmap showing the hierarchical cluster of IgG and IgA autoantibodies significantly different when comparing young versus elderly controls or young versus elderly of each COVID-19 cohort. The colors blue and light yellow indicate autoantibodies with or without significant differences between the young versus elderly in each group. **b)** Box plots display changes in the autoantibody levels by age (young < 60 and elderly ≥ 60). Each box plot shows the median with first and third interquartile range (IQR), whiskers representing minimum and maximum values within IQR, and individual data points. Significance was determined using two-sided Wilcoxon rank-sum tests and is indicated by asterisks (* p ≤ 0.05, ** p ≤ 0.01, *** p ≤ 0.001, and **** p ≤ 0.0001). control-young (*n* = 65) and control-elderly (*n* = 12); mild-young (*n* = 70), mild-elderly (*n* = 4), moderate-young (*n* = 45), moderate-elderly (*n* = 20), severe-young (*n* = 20), severe-elderly (*n* = 12) COVID-19 patients. All autoantibodies with significant differences between at least two groups are shown in **Supplementary Fig. 7**.

Of note, there were only a few (IgG autoantibodies targeting heparin and LSPC; SARS-CoV-2 antibodies; IgA autoantibodies targeting cardiolipin, DEC, ICA, and tyrosinase) age-dependent significant differences in patients with anosmia versus no anosmia regarding the top 10 autoantibody predictors of COVID-19 severity (**Supplementary Fig. 8**). Furthermore, we asked whether sex also impacts the levels of autoantibodies during SARS-Co-2 infections. IgG anti-cardiolipin-aab was elevated in males from all three disease groups (but only significantly in mild and moderate COVID-19 cohorts) when compared to females. Except for this, there were no significant sex differences in the levels of the top 10 IgG and IgA autoantibody predictors of COVID-19 severity (**Supplementary Fig. 9**).

## DISCUSSION

This work sheds light on the immunopathogenesis of COVID-19 demonstrating the presence of increased levels of IgG and IgA autoantibodies linked to diverse number of autoimmune diseases. Among them, are autoantibodies not yet reported in COVID-19 patients such as those targeting neuronal antigens (e.g., α-synuclein, acetylcholine receptor, myeloid-β peptide, β-catenin, brain-derived neurotrophic factor, cerebellar antigen, chondroitin sulphate, dopamine receptors [D1 and D2], enteric nerve, ganglioside, glutamic acid decarboxylase, myelin basic protein, myelin oligodendrocyte glycoprotein, neurofilament proteins, NMDA receptor, rabaptin-5, somatotropin, S100B, tau protein, and transglutaminases 6) and non-neuronal antigens (e.g., α-enolase, α-myosin, claudin-5 and -7, collagen, desmoglein-E-cadherin 1, epithelial sodium channel α, fibulin, fibrinogen, human epidermal keratin, insulin receptor, islet cell antigen, occluding, platelet glycoprotein, transglutaminases 2 and 3, tyrosinase, and zonulin).

Importantly, autoantibodies correlate with clinical features of SARS-CoV-2 infection such as disease severity and anosmia. Besides expanding the spectrum of autoantibodies associated with COVID-19 (e.g., cardiolipin^72^, dsDNA^72^, epithelial cell antigen^73^, heparin^74^, human RO60^75^, liver microsomal antigen^40^, lung surfactant protein C)^76^, our data also indicate that both disease severity and anosmia disrupt the physiological IgG and IgA autoantibody signatures. These findings are consistent with the notion that autoantibodies are natural components of human physiology and become dysregulated under pathological conditions^19,20,77,78^. Further supporting the concept that COVID-19 disrupts antibody physiology, anti-SARS-CoV-2 antibody levels accompany the levels of autoantibodies. Those antibodies targeting self- and non-self-antigens, along with age, are predictors of disease severity as classified by random forest modeling. Viral-induced autoimmune diseases are due to multiple mechanisms such as molecular mimicry^79^, epitope spreading^80^, and bystander activation, all of which could possibly be involved in SARS-CoV-2 induced autoimmunity. Indeed, evidence has been provided for strong, chronic inflammation promoting the release of self-antigens and high cytokine levels activating bystander T-cells^81^, and for molecular mimicry^82–84^, all of which have been associated with severe COVID-19. Molecular mimicry has been explored by assessing the SARS-CoV-2 proteome, which revealed the existence of 21 viral peptides that show at least 90% homology with human proteins known to be involved in autoimmune diseases such as multiple sclerosis and rheumatoid arthritis^82^. Furthermore, it has been shown that monoclonal antibodies generated against SARS-CoV-2 proteins reacted with 28 out of 55 tested autoantigens^70-71^. These observations may explain why the multiple serum autoantibodies causative to autoimmune diseases dysregulate their levels, perturbing their physiologic function. These findings are in line with reports from ours^20^ and other groups^10,40^ demonstrating that changes in autoantibody signatures affect physiologic and pathologicl immune homeostasis^20^.

Of note, the random forest model suggests that anti-SARS-CoV-2 antibodies and autoantibodies, together with the major risk factor age^71^, are important predictors of COVID-19 severity. Our finding that the highest levels of anti-SARS-CoV-2 antibodies are achieved by patients with severe COVID-19 is in agreement with previous reports demonstrating a similar pattern of humoral immune response in critically ill COVID-19 patients^75,85^. In addition, it was shown that the levels of anti-SARS-CoV-2 antibodies accompany longitudinally with the dysregulation of autoantibody levels^40^. While we found no age effect on the levels of anti-SARS-CoV-2 antibodies, severe SARS-CoV-2 infection induces higher autoantibody levels in elderly patients compared with young patients. Thus, our data point to novel mechanisms involved in the risk intersection of immunosenescence and COVID-19^71^, suggesting that SARS-CoV-2 infection induces a broader loss of self-tolerance than previously thought, particularly in elderly patients. In this context, several age-associated factors such as chronic inflammation in ageing (inflammageing^86^) might promote the production of autoantibodiesas well as the tendency to naturally progress to immune dysregulation of innate^87^ and adaptive^88^ immune cells. Therefore, our data support the idea that the induction of high levels of serum autoantibodies in elderly COVID-19 patients contribute to the increased risk for negative outcomes in older persons^89^.

Because autoantibodies are strong predictors of COVID-19 severity, our findings expand the number of autoantibodies that qualify as biomarkers for the severity of SARS-CoV-2 infection. Among them are those targeting molecules involved in neurological functions (e.g.,cognition, learning, memory, synaptic transmission) and glucose metabolism. For instance, while anti-cardiolipin antibodies were previously associated with the development of hyperinflammatory syndromes^90^, the high levels of autoantibodies targeting neuronal-associated molecules could provide clues why the respiratory symptoms of COVID-19 patients are often accompanied by short- and long-term neuropsychiatric symptoms and brain sequelae^68^. It has been convincingly shown that SARS-CoV-2 is able enter the brain by crossing the blood-brain barrier (BBB) because inflammatory cytokines induce BBB instability^68,91,92^. Our data indicate that the array of autoantibodies targeting neuronal molecules is an additional molecular layer that could contribute to the neurological manifestations^66,67^ occuring in COVID-19 patients.

There are limitations of our worlk that need consideration. For instance, our study did not have longitudinal data to analyze the pharmacokinetics of the IgG and IgA autoantibodies, from disease onset to convalescence or post-acute COVID-19 syndrome. Our study cohort did not include asymptomatic patients. Moreover, we cannot exclude the possibility that at least some of our patients had high levels of autoantibodies prior to SARS-CoV-2 infection, or that autoantibodies to some antigens (e.g., heparin) could have been induced by anticoagulant therapy with heparin. In addition, we did not assess alterations in the number of circulating B lymphocytes and whether this could explain the higher serum levels of autoantibodies. In addition, future studies are required to clarify the role of virus and host genetics in the production of autoantibodies. On the other hand, our work raises new questions such as whether the dysregulated levels of autoantibodies remain after COVID-19 remission and these levels are in patients with post-COVID-19 syndrome.

Taken together, our work provides a comprehensive view of the spectrum of autoantibodies linked with autoimmune diseases that are induced by SARS-CoV-2 infection. This work maps the intersection of COVID-19 and autoimmunity^93–95^, demonstrating the dysregulation of multiple autoantibodies that are linked to autoimmune diseases during SARS-CoV-2 infections, and the altered correlation signatures according to disease severity and anosmia. The data presented suggest a gradual involvement of autoantibodies in the severity of COVID-19. We identified several new clinically relevant autoantibodies that can be used as biomarkers that predict COVID-19 severity and to open avenues to develop new treatment strategies.

## Data Availability

All data present in this manuscript are provided as Supplementary files. All R packages used in this manuscript are available at the following link.

https://github.com/gabrielacbaiocchi/Autoantibodies-linked-to-autoimmune-diseases-associate-with-COVID-19-outcomes.git

## Acknowledgments

We acknowledge the patients for participation in this study. We would like to acknowledge the contributions of Lev Rochel Bikur Cholim of Lakewood (led by Rabbi Yehuda Kasirer and Mrs. Leeba Prager) and their hundreds of volunteers who participated in collecting samples for this research through the MITZVA Cohort. We thank Immunosciences and Cyrex Laboratories for financial support and INOVA Diagnostics for providing their diagnostic ELISA kits for autoimmunity at a very discounted rate Furthermore, we would like to thank Vilma Samayoa, David Cisneros, Roberto Melgar, Dana Ashley Hill and Amanda Thornton for technical assistance.. We also thank the São Paulo Research Foundation (FAPESP grants: 2020/07972-1 to GCB; 2020/09146-1 to PPF; 2020/11710-2 to DRP, 2018/18886-9, 2020/01688-0, and 2020/07069-0 to OCM), and the Coordination for the Improvement of Higher Education Personnel (CAPES) Financial Code 001 (grant to ISF) for financial support.

## Data availability

All data present in this manuscript are provided as Supplementary files. All R packages used in this manuscript are are available at the following link: https://github.com/gabrielacbaiocchi/Autoantibodies-linked-to-autoimmune-diseases-associate-with-COVID-19-outcomes.git

## Author contributions

YS, AZR, IZ, AV, and EV conceived the project; YS, AZR, IZ, and GH designed the study. JIS, AZR, IZ, and MTL diagnosed, recruited or followed-up the patients. AV and EV performed the experiments. AZR, DAH, AT, GH, JZ, JIS, AZR, IZ, MTL, EV, YBL, HA, and YS coordinated the serum collection and databank. GCB, AHCM, ISF, DLMF, DRP, PPF, MK, RDV, and OCM performed data and bioinformatics analyses. GCB, AV, LMG, HDO, and OCM wrote the manuscript. JS, HH, YO, AZR, VLGC, NOSC, RDV, IZ, GCB, AV, LFS, LMG, HDO, and OCM edited the manuscript; GCB, AV, AZR, YO, LFS, IZ, AHCM, EV, DAH, AT, LMG, YBL, JIS, JZ, NOSC, VLGC, HH, GR, HA, OCM, and YS, provided scientific insights.

## Competing interests

The authors declare no competing interests.

## Notes

### Competing Interest Statement

The authors have declared no competing interest.

### Author Declarations

APPROVED BY INTEGREVIEW IRB NOVEMBER 21, 2020

