## Supplementary Figures 1 to 9 for "Autoantibodies linked to autoimmune diseases associate with COVID-19 outcomes"

Supplementary Figures 1-9

Supplementary Tables S1-S6  provided as Supplementary Data file

**Suppl. Figure 1**

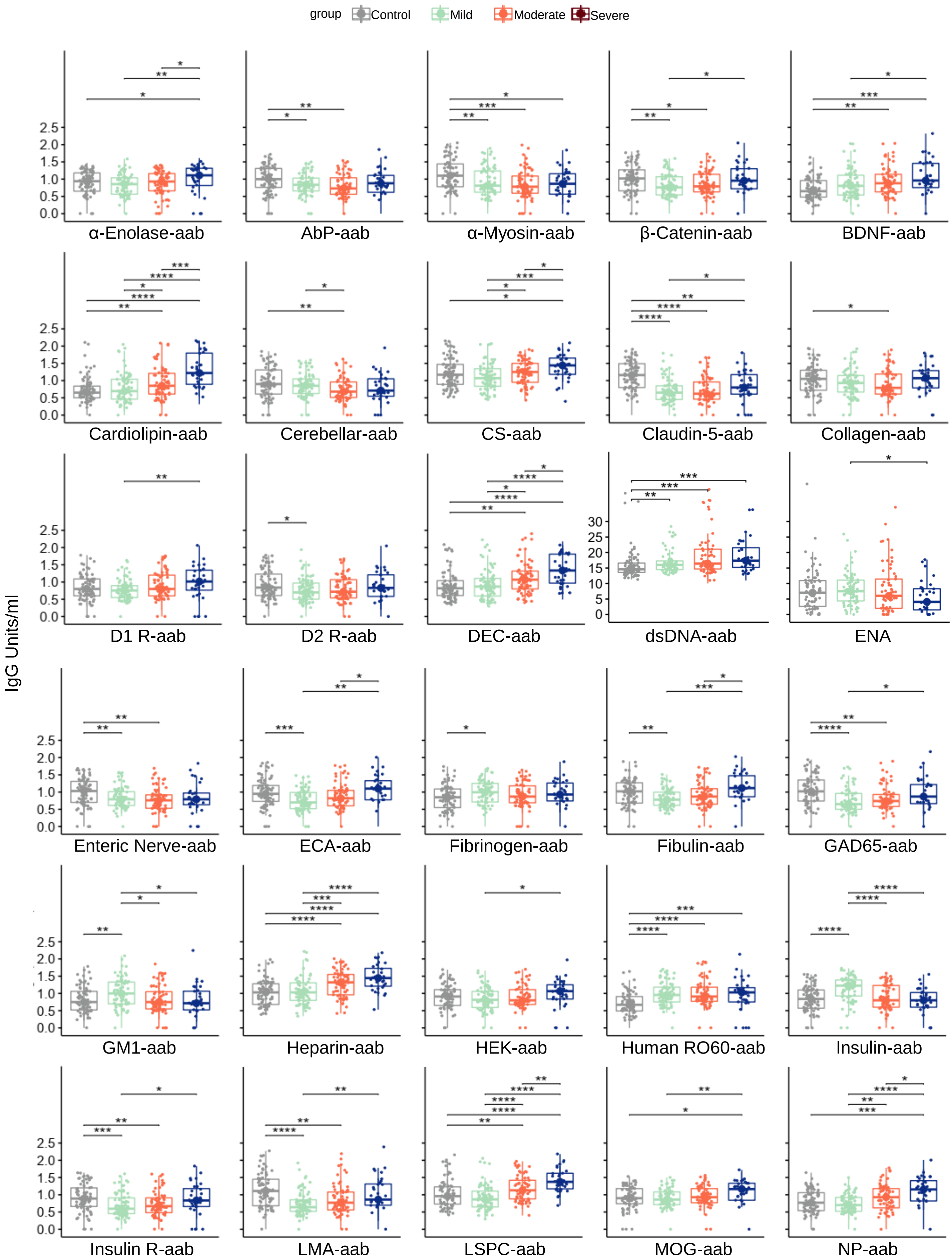

#### Suppl. Figure 1

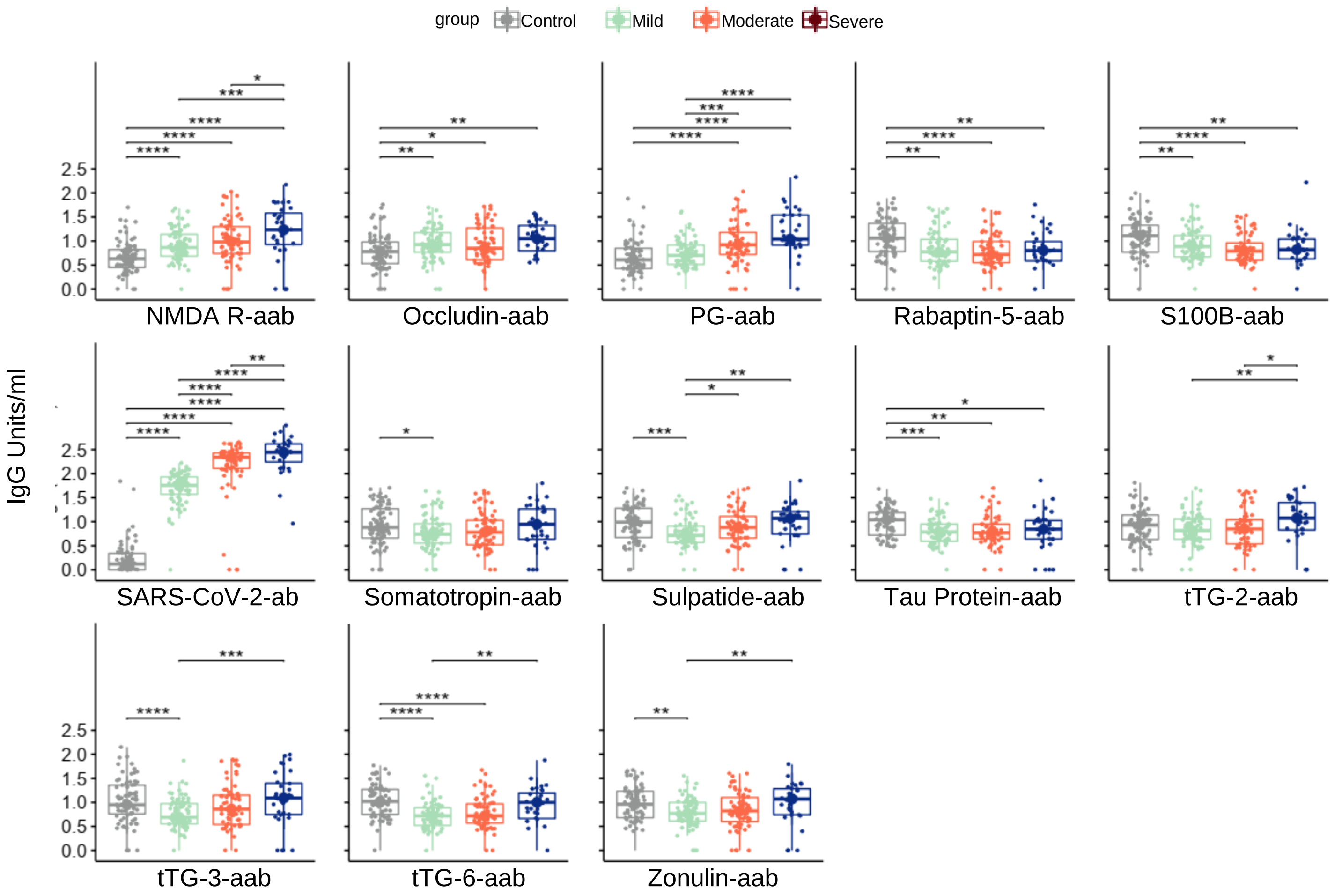

### Suppl. Figure 2

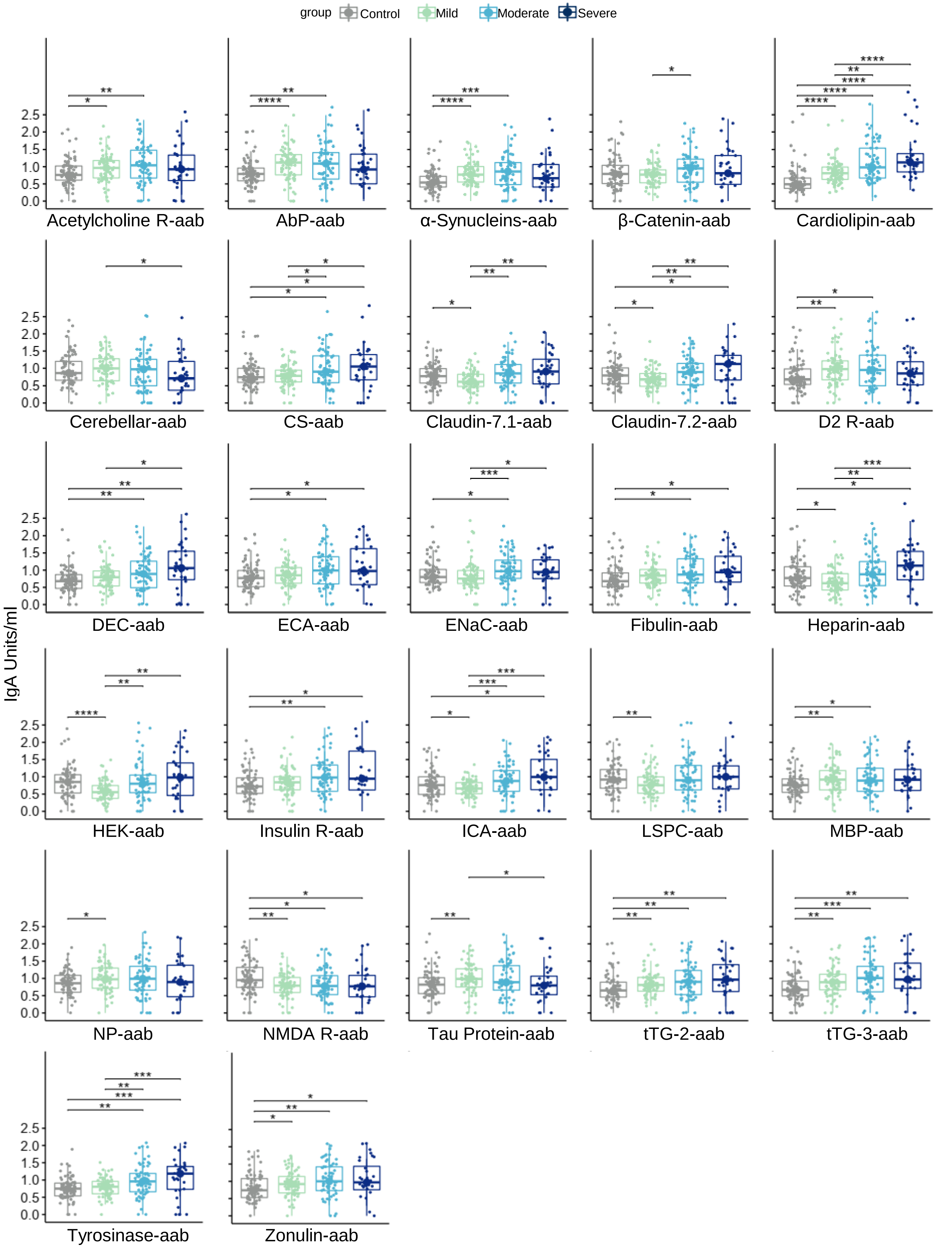

**Suppl. Figure 3**

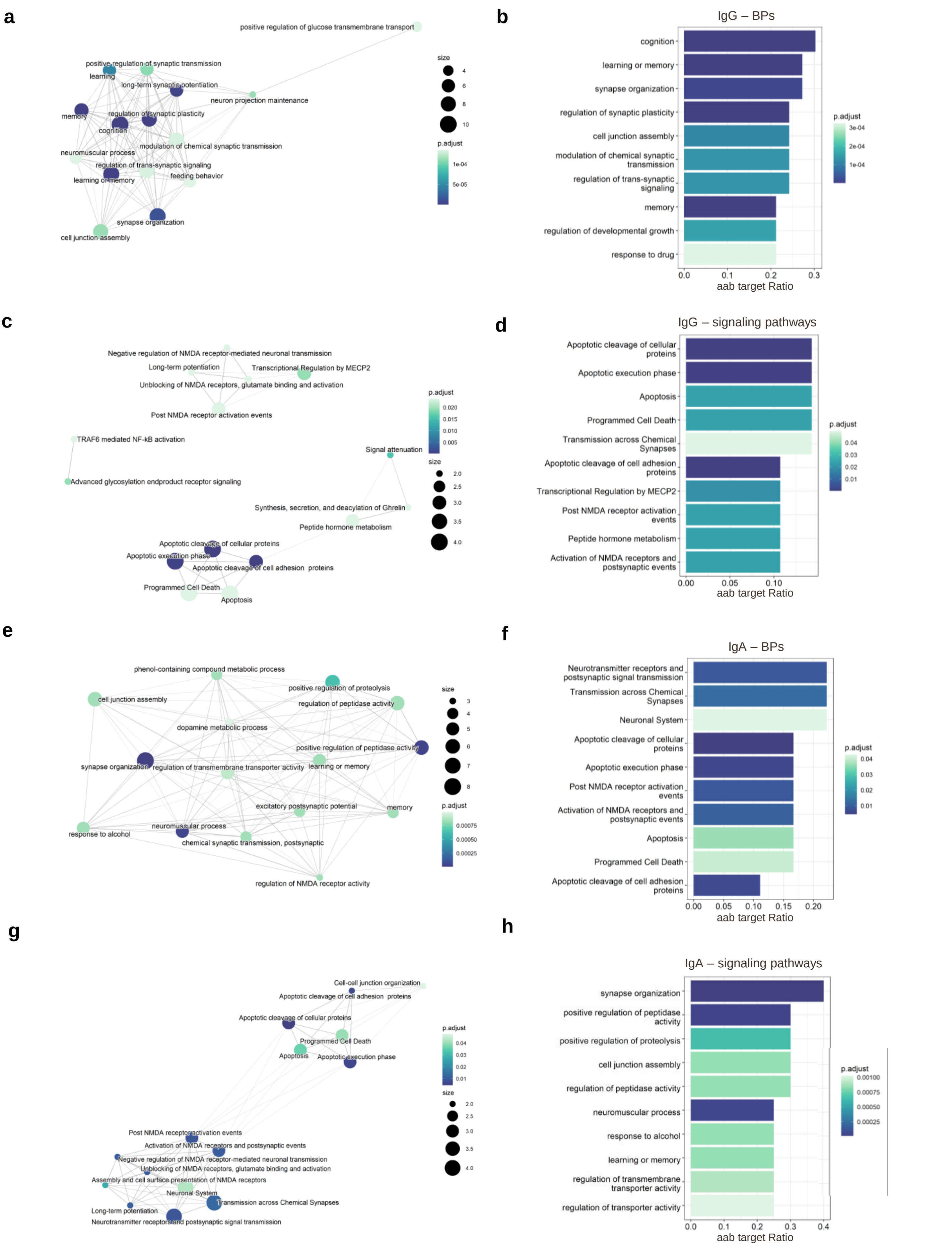

**a**

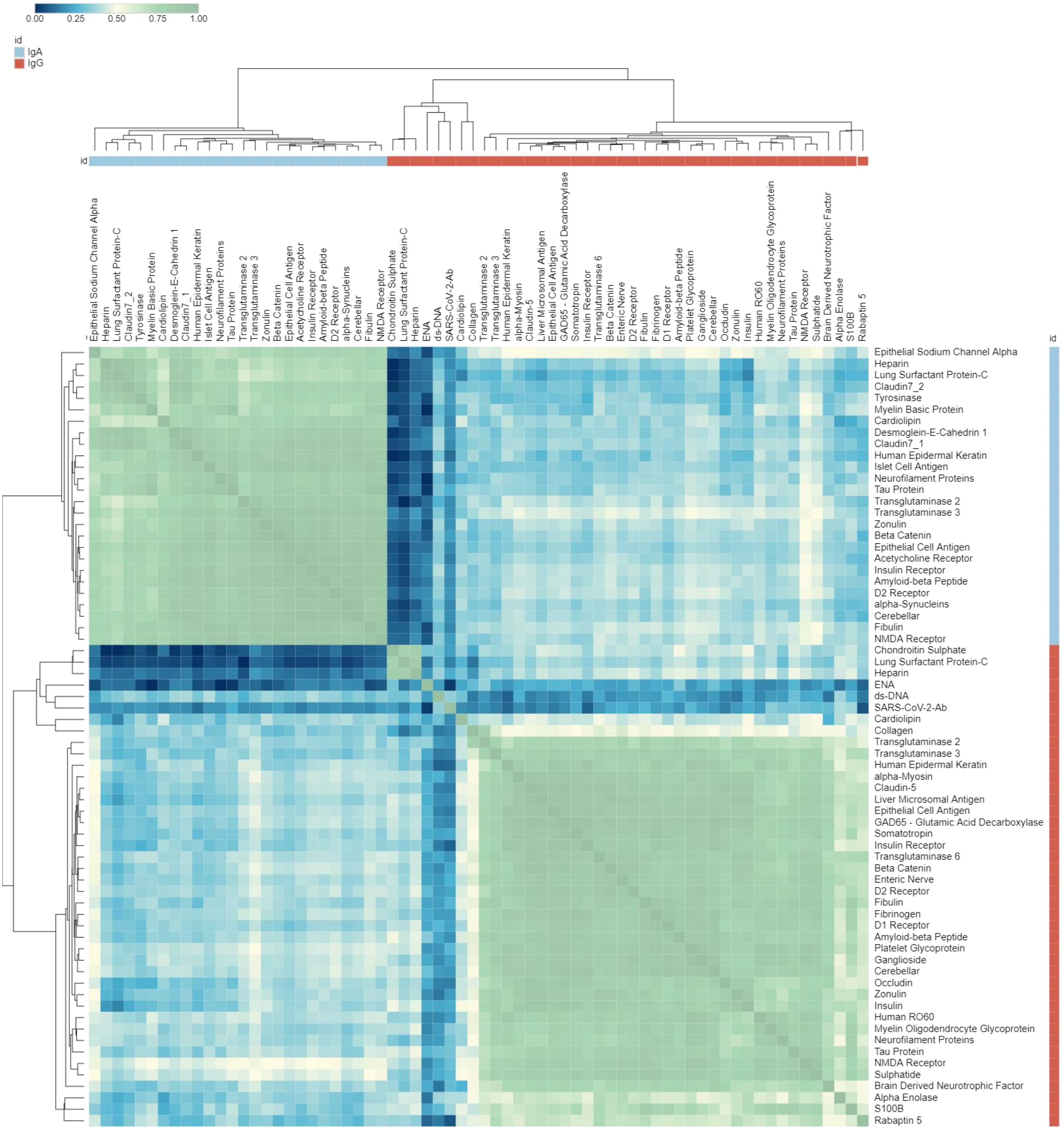
0.00
0.25
0.50
0.75
1.00
id
IgA
IgG
Epithelial Sodium Channel Alpha
Heparin
Lung Surfactant Protein-C
Claudin7_2
Tyrosinase
Myelin Basic Protein
Cardiolipin
Desmoglein-E-Cahedrin 1
Claudin7_1
Human Epidermal Keratin
Islet Cell Antigen
Neurofilament Proteins
Tau Protein
Transglutaminase 2
Transglutaminase 3
Zonulin
Beta Catenin
Epithelial Cell Antigen
Acetycholine Receptor
Insulin Receptor
Amyloid-beta Peptide
D2 Receptor
alpha-Synucleins
Cerebellar
Fibulin
NMDA Receptor
Chondroitin Sulphate
Lung Surfactant Protein-C
Heparin
ENA
ds-DNA
SARS-CoV-2-Ab
Cardiolipin
Collagen
Transglutaminase 2
Transglutaminase 3
Human Epidermal Keratin
alpha-Myosin
Claudin-5
Liver Microsomal Antigen
Epithelial Cell Antigen
GAD65 - Glutamic Acid Decarboxylase
Somatotropin
Insulin Receptor
Transglutaminase 6
Beta Catenin
Enteric Nerve
D2 Receptor
Fibulin
Fibrinogen
D1 Receptor
Amyloid-beta Peptide
Platelet Glycoprotein
Ganglioside
Cerebellar
Occludin
Zonulin
Insulin
Human RO60
Myelin Oligodendrocyte Glycoprotein
Neurofilament Proteins
Tau Protein
NMDA Receptor
Sulphatide
Brain Derived Neurotrophic Factor
Alpha Enolase
S100B
Rabaptin 5

Control

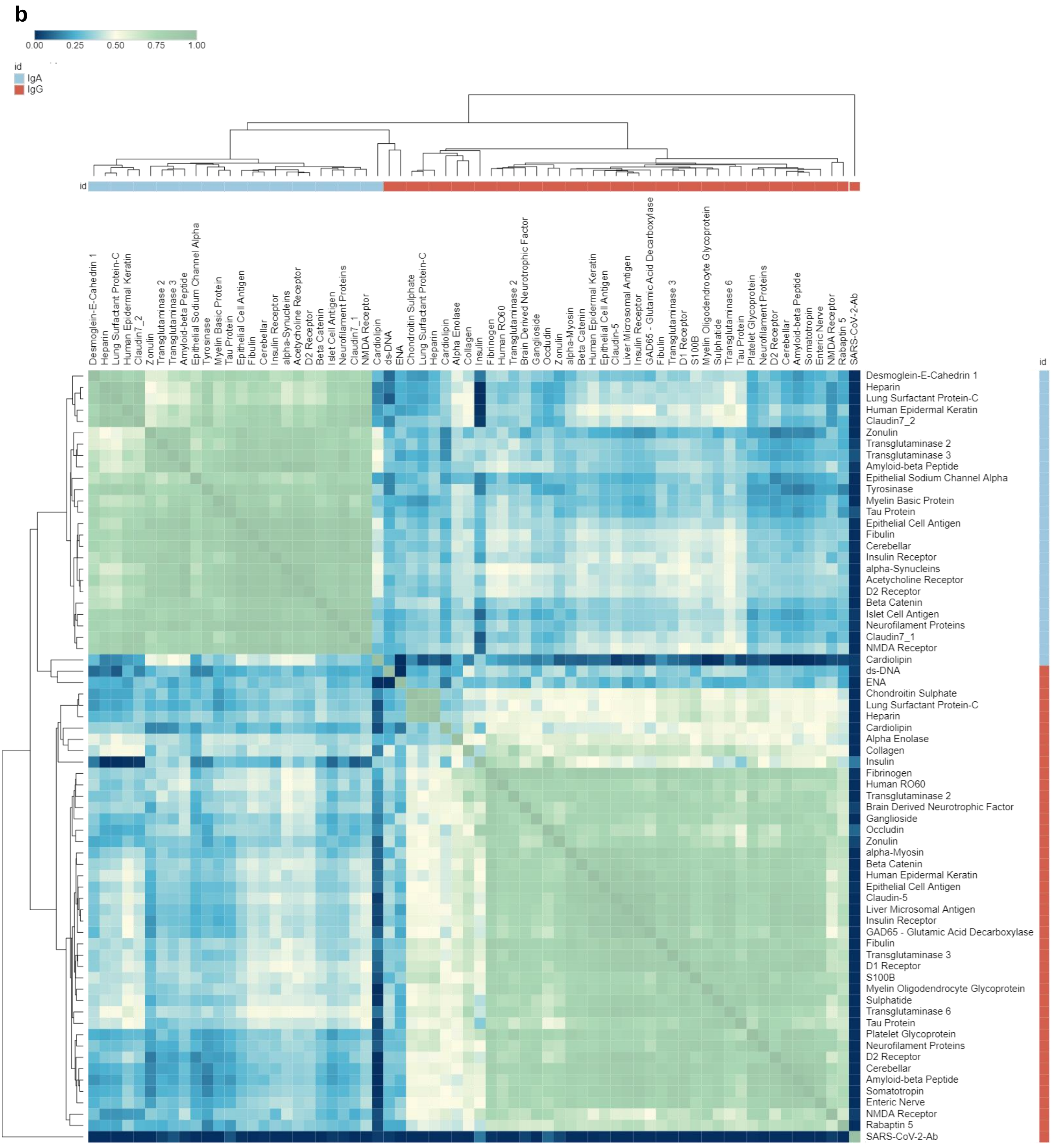


Mild COVID-19

**c**

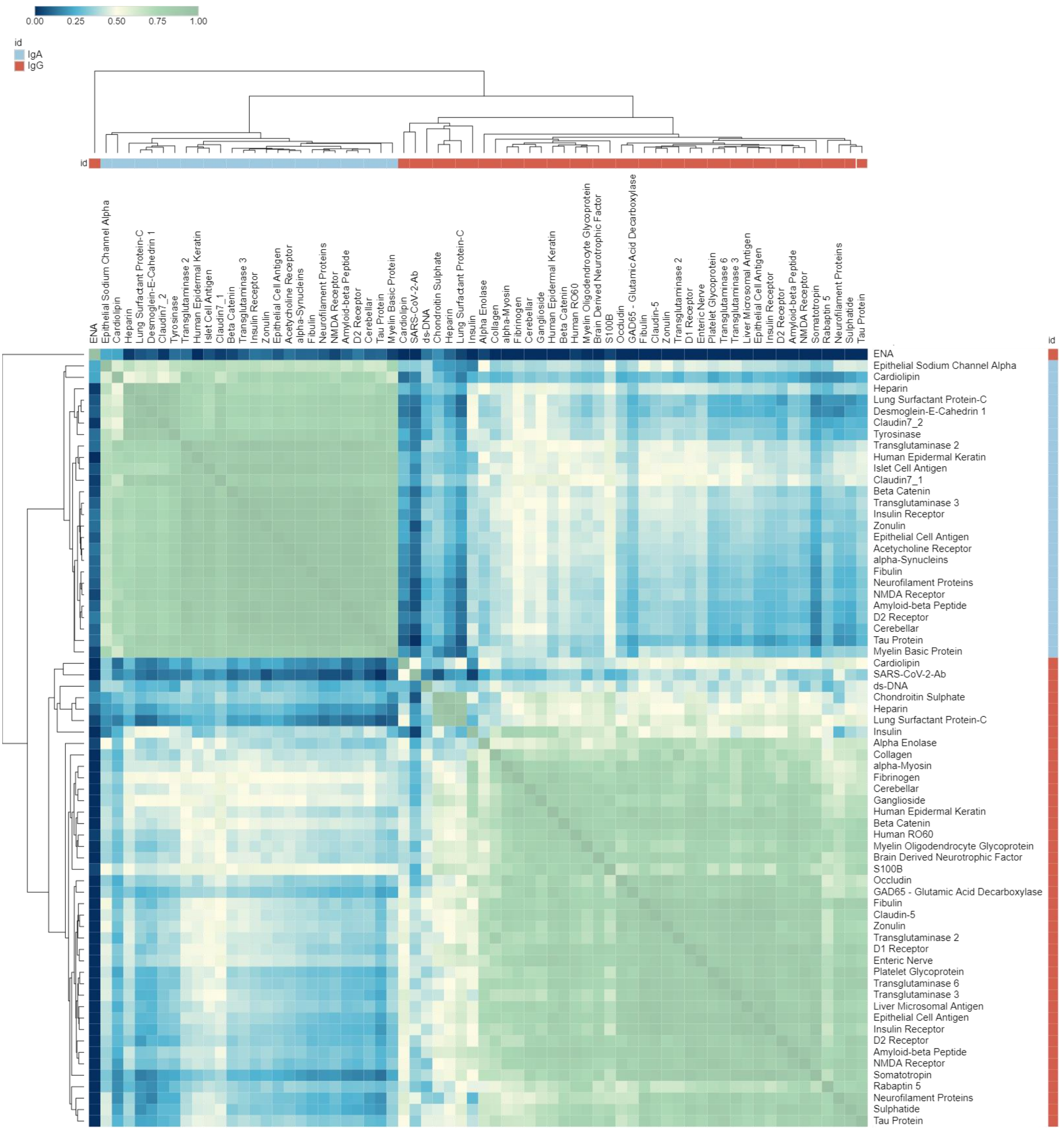


Moderate COVID-19

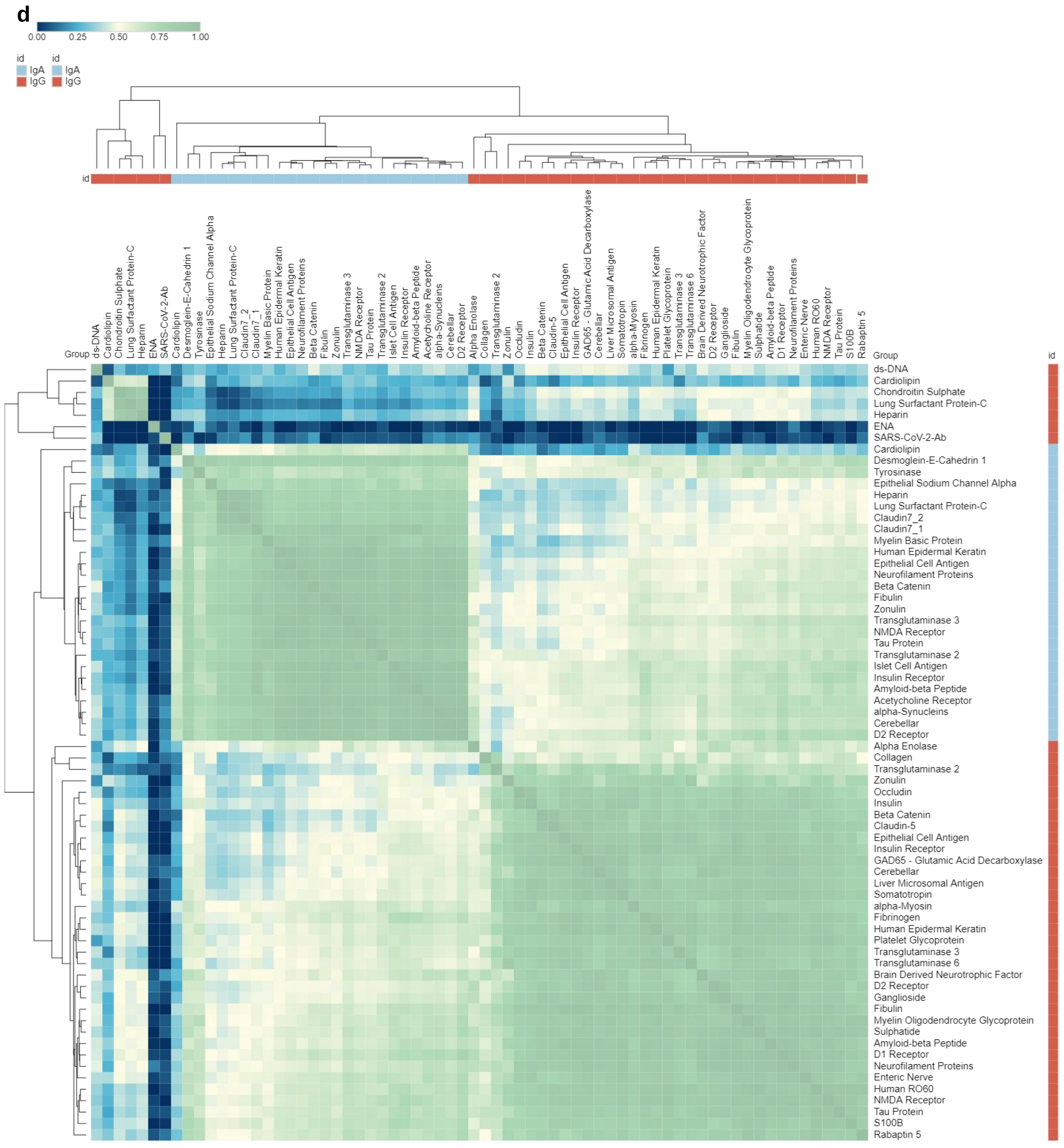


Severe COVID-19

**Cont. Suppl. Figure 4**

**e**

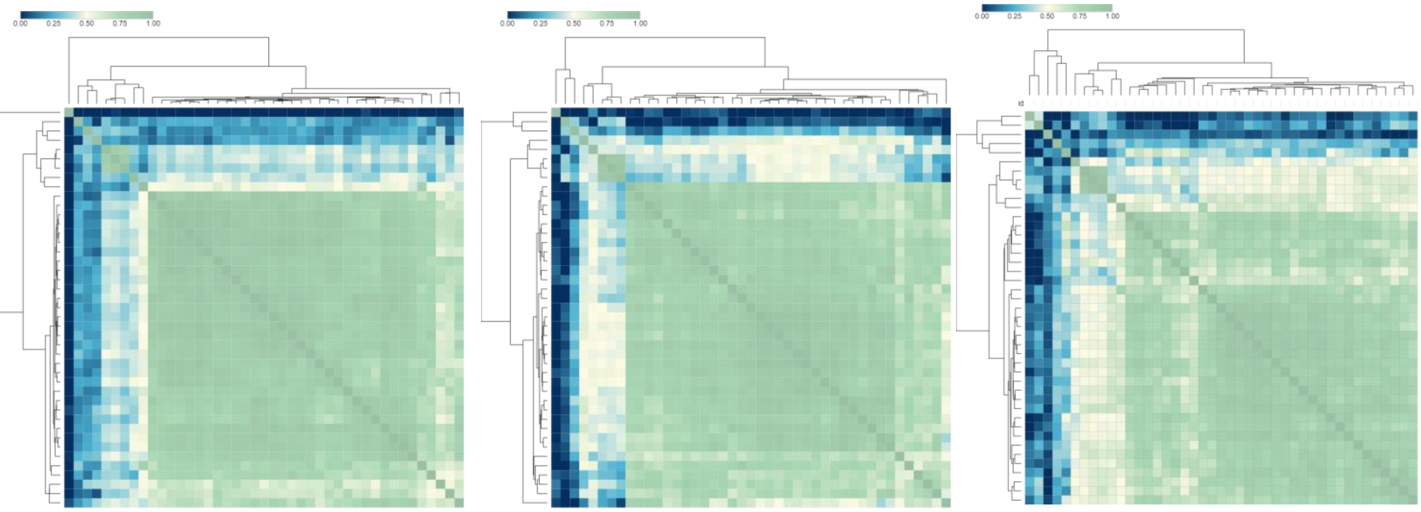


Control (IgG) | COVID-19 No Anosmia (IgG) | COVID-19 Anosmia (IgG)

**Suppl. Figure 5**

**a**

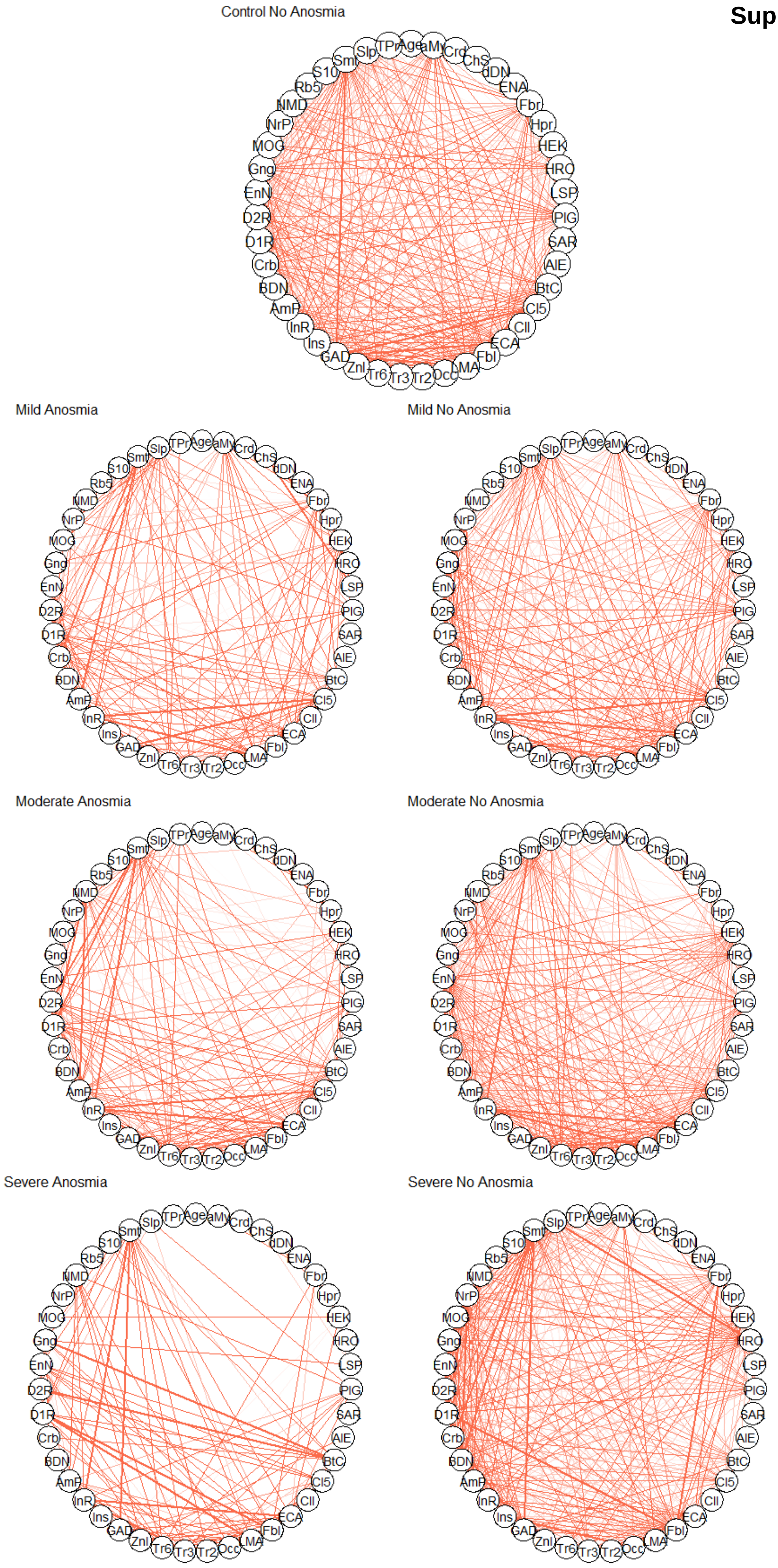

**Cont. Suppl. Figure 5**

**b**

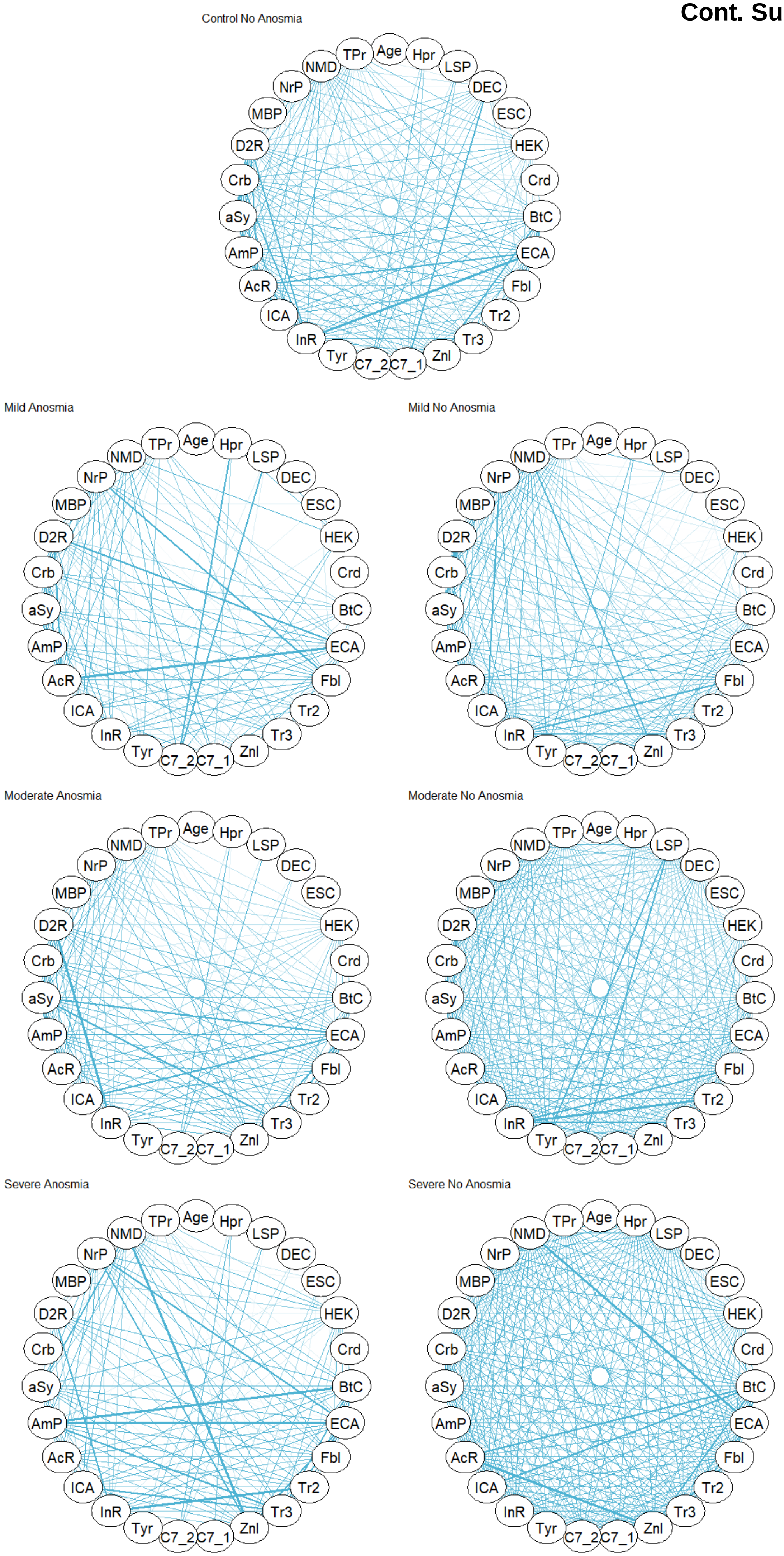

### Suppl. Figure 6

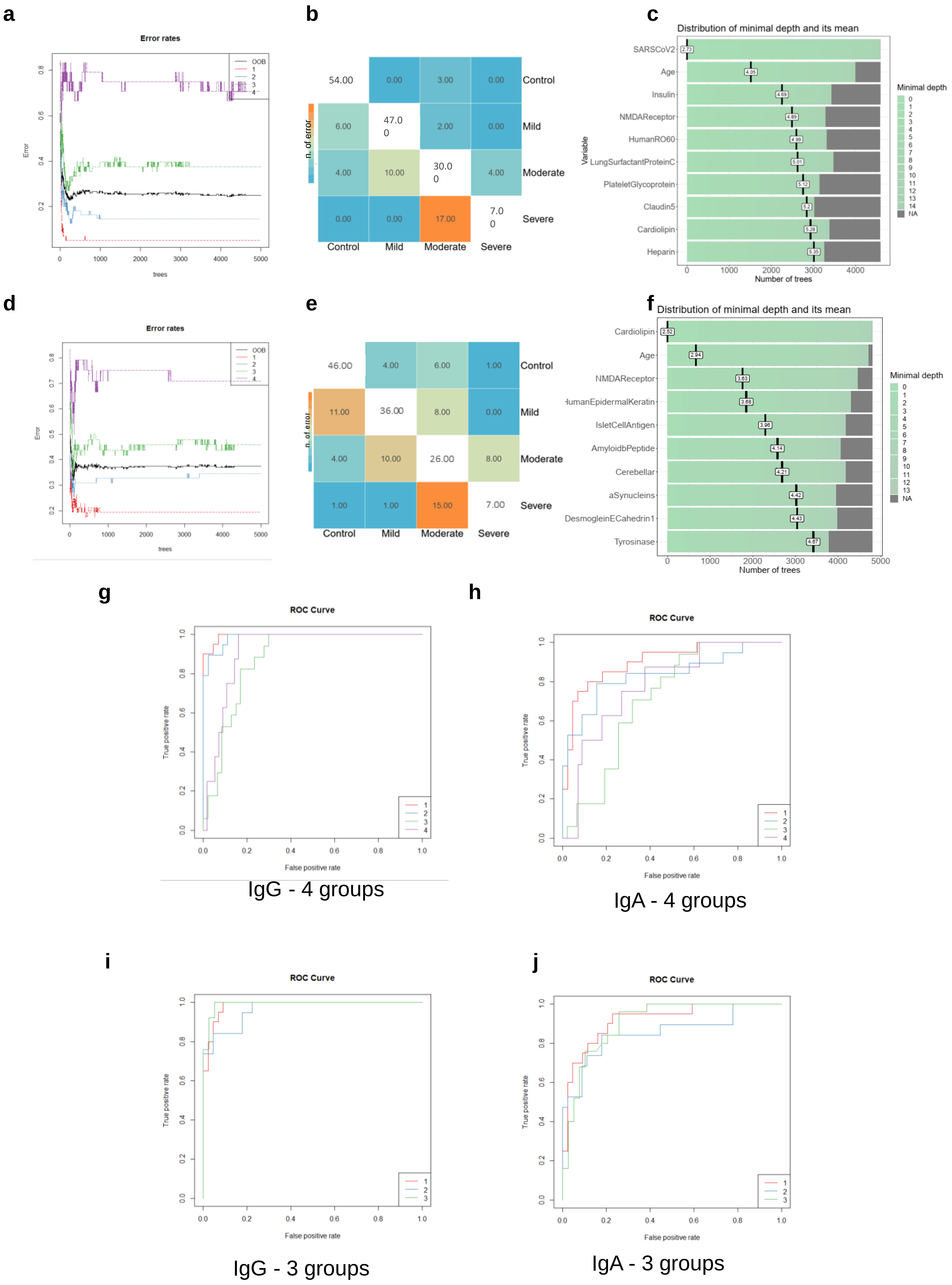

### Suppl. Figure 7

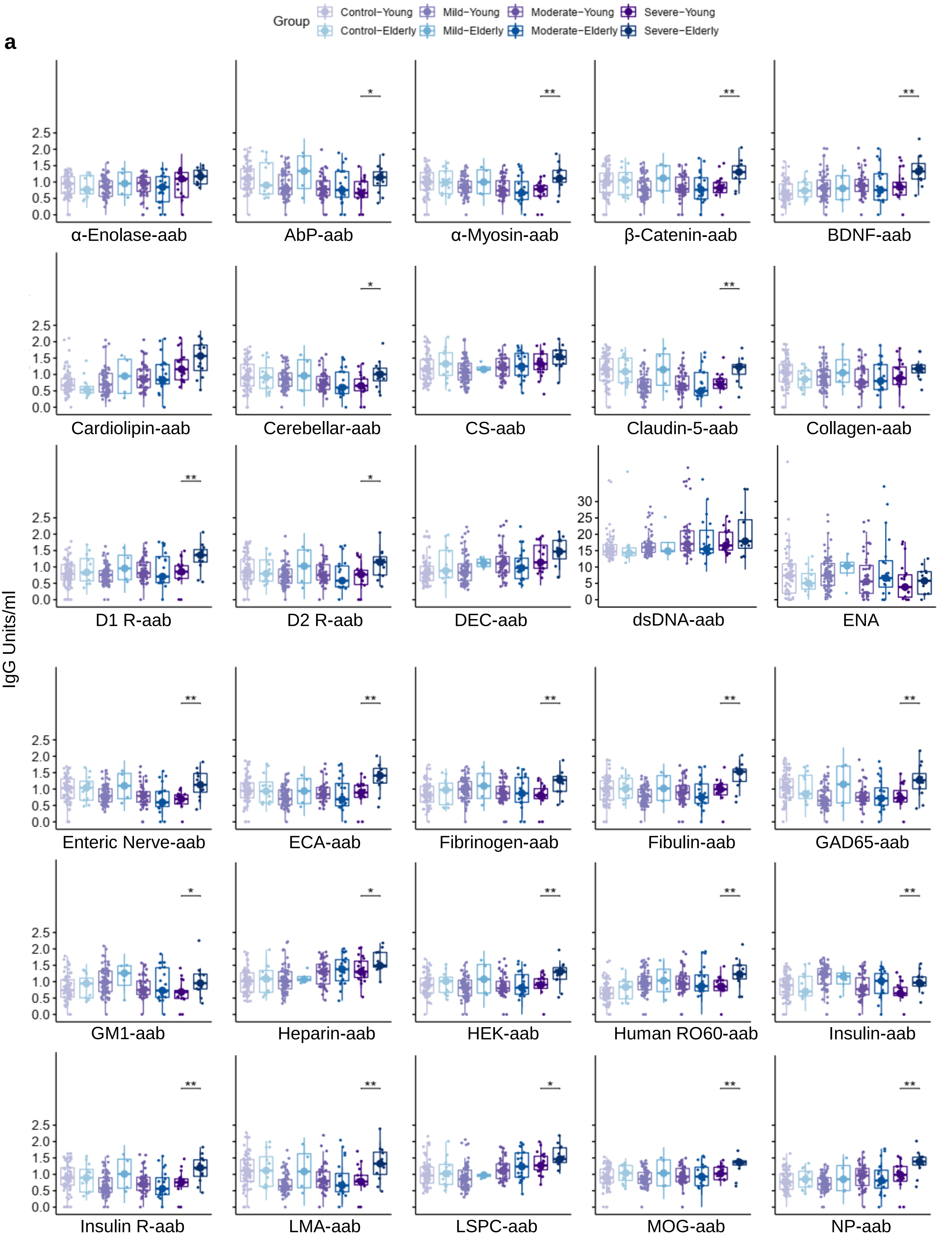

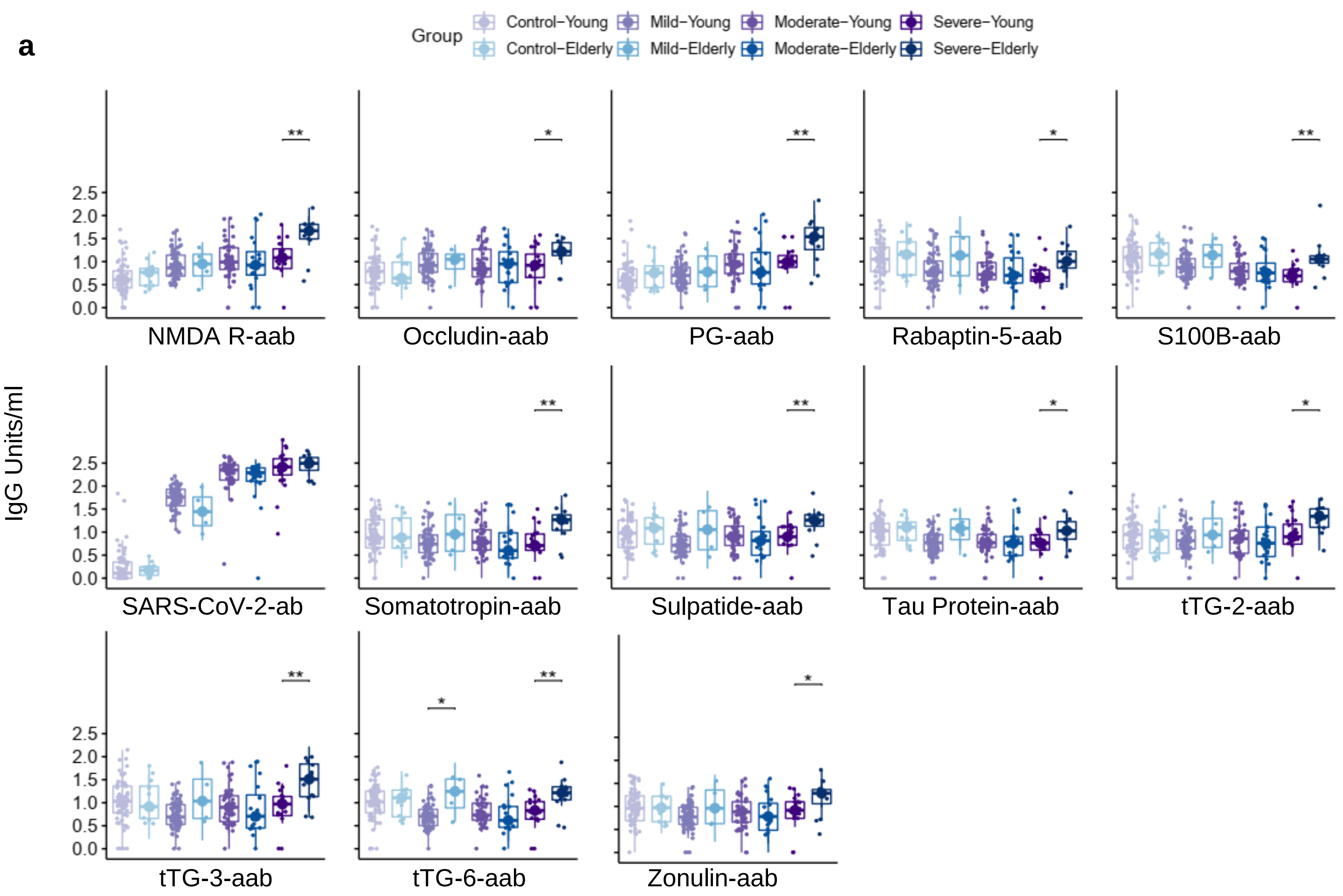
a
Group
Control−Young
Control−Elderly
Mild−Young
Mild−Elderly
Moderate−Young
Moderate−Elderly
Severe−Young
Severe−Elderly
IgG Units/ml
NMDA R-aab
Occludin-aab
PG-aab
Rabaptin-5-aab
S100B-aab
SARS-CoV-2-ab
Somatotropin-aab
Sulpatide-aab
Tau Protein-aab
tTG-2-aab
tTG-3-aab
tTG-6-aab
Zonulin-aab

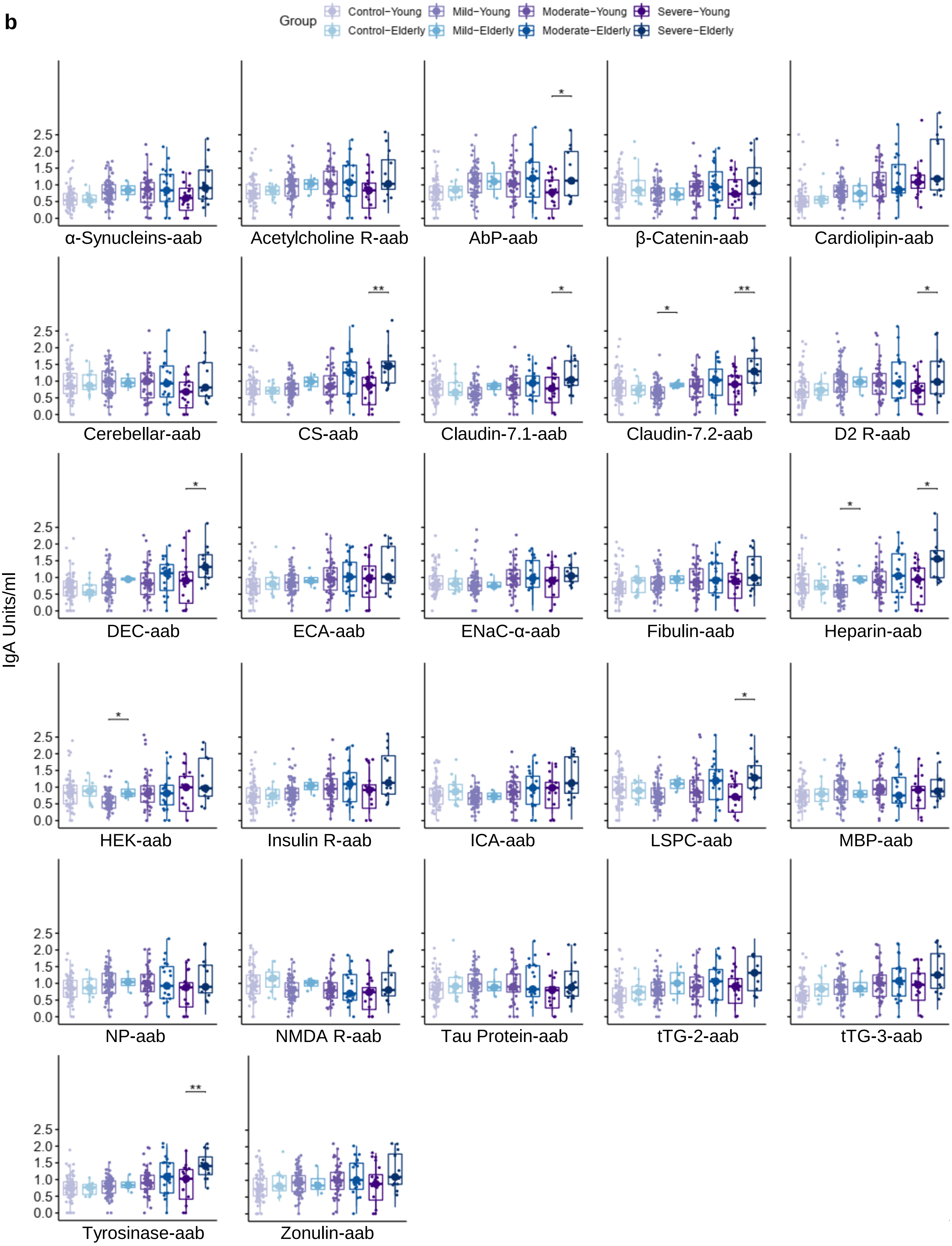
b
Group
Control−Young
Control−Elderly
Mild−Young
Mild−Elderly
Moderate−Young
Moderate−Elderly
Severe−Young
Severe−Elderly
IgA Units/ml
α-Synucleins-aab
Acetylcholine R-aab
AbP-aab
β-Catenin-aab
Cardiolipin-aab
Cerebellar-aab
CS-aab
Claudin-7.1-aab
Claudin-7.2-aab
D2 R-aab
DEC-aab
ECA-aab
ENaC-α-aab
Fibulin-aab
Heparin-aab
HEK-aab
Insulin R-aab
ICA-aab
LSPC-aab
MBP-aab
NP-aab
NMDA R-aab
Tau Protein-aab
tTG-2-aab
tTG-3-aab
Tyrosinase-aab
Zonulin-aab

**Suppl. Figure 8**

a

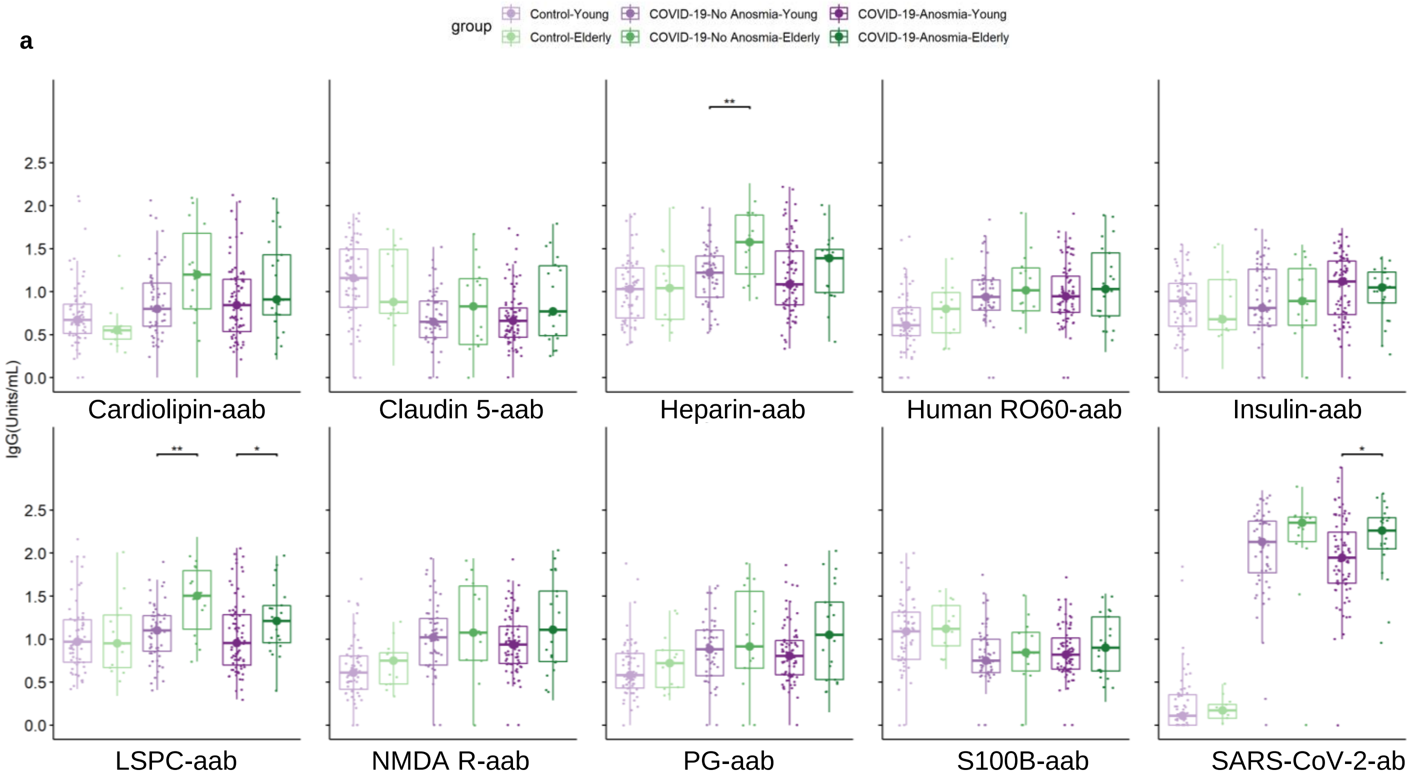


b

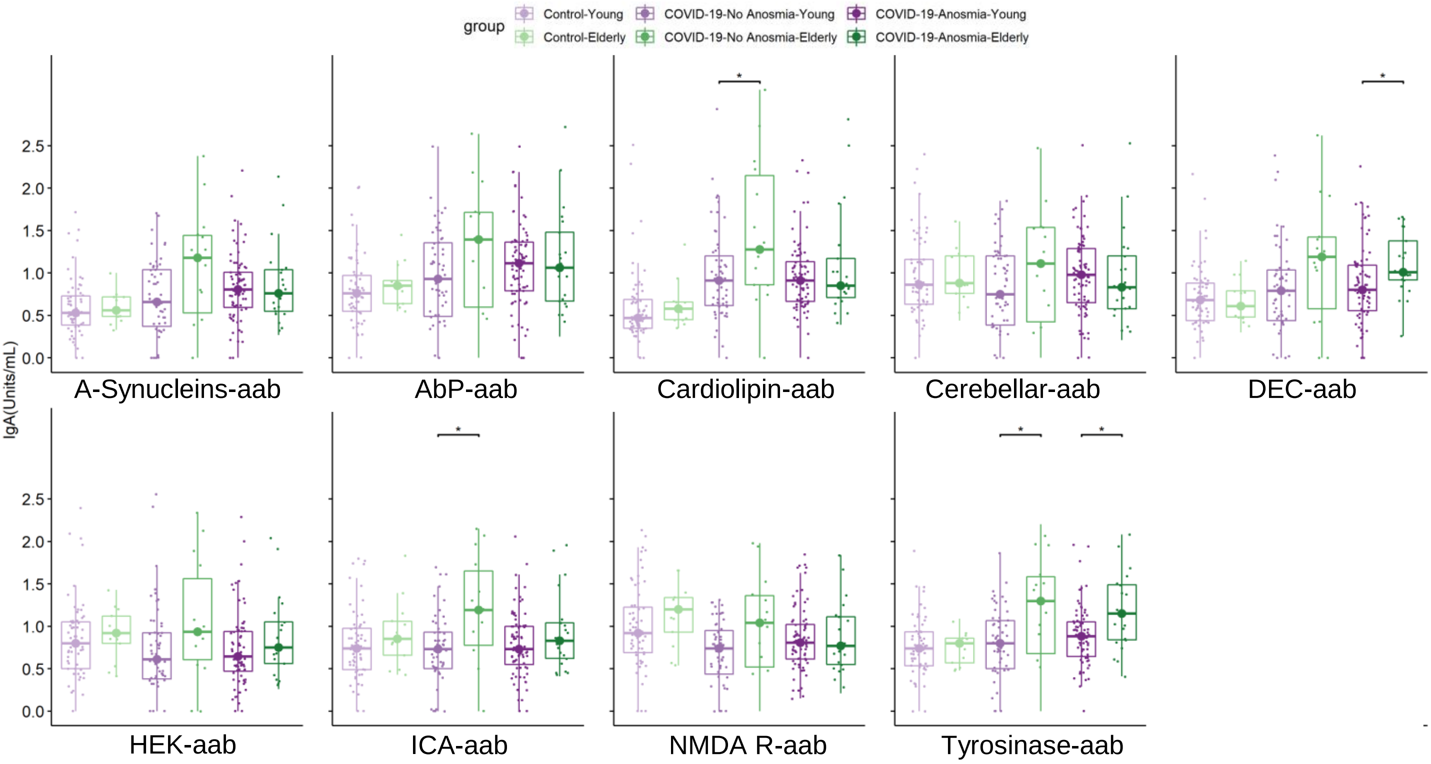

**Suppl. Figure 9**

a

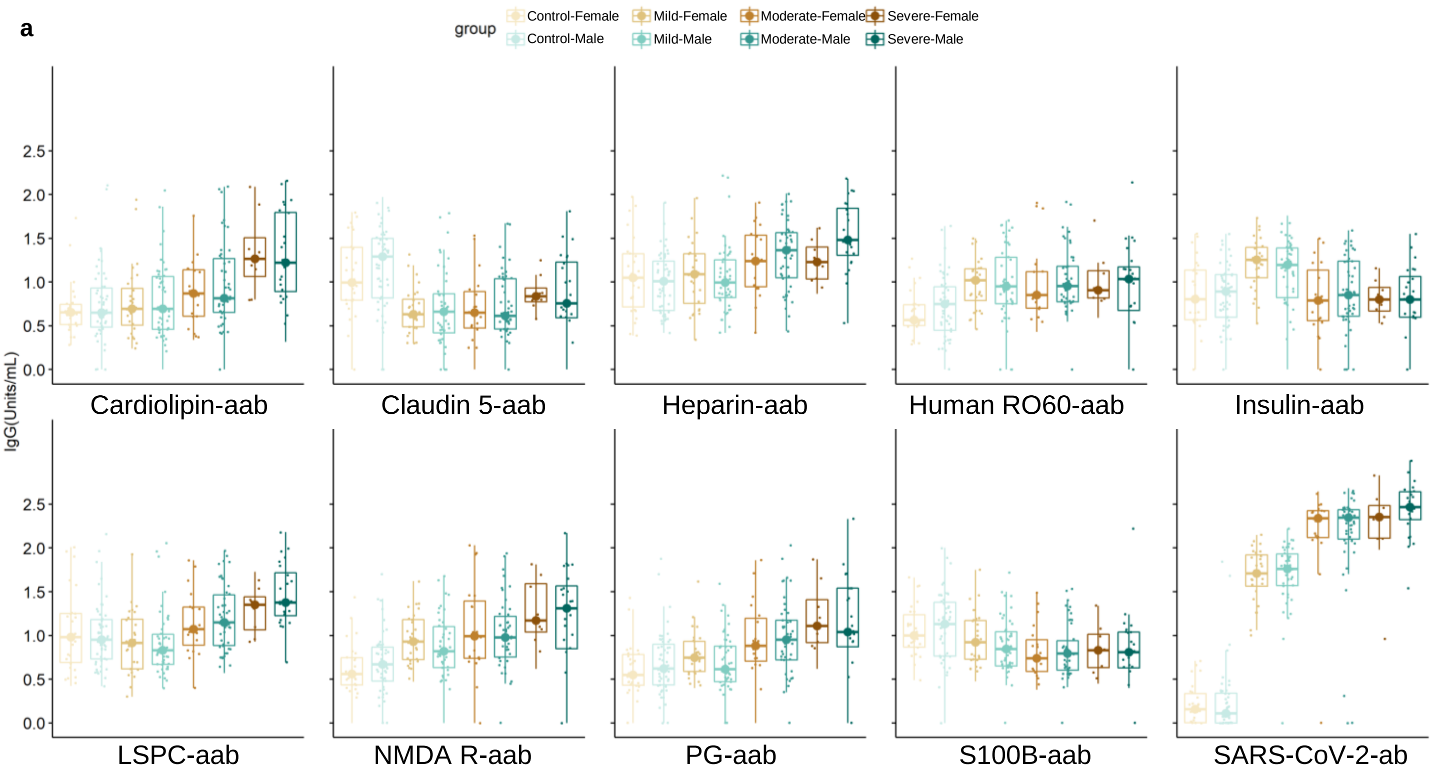


b

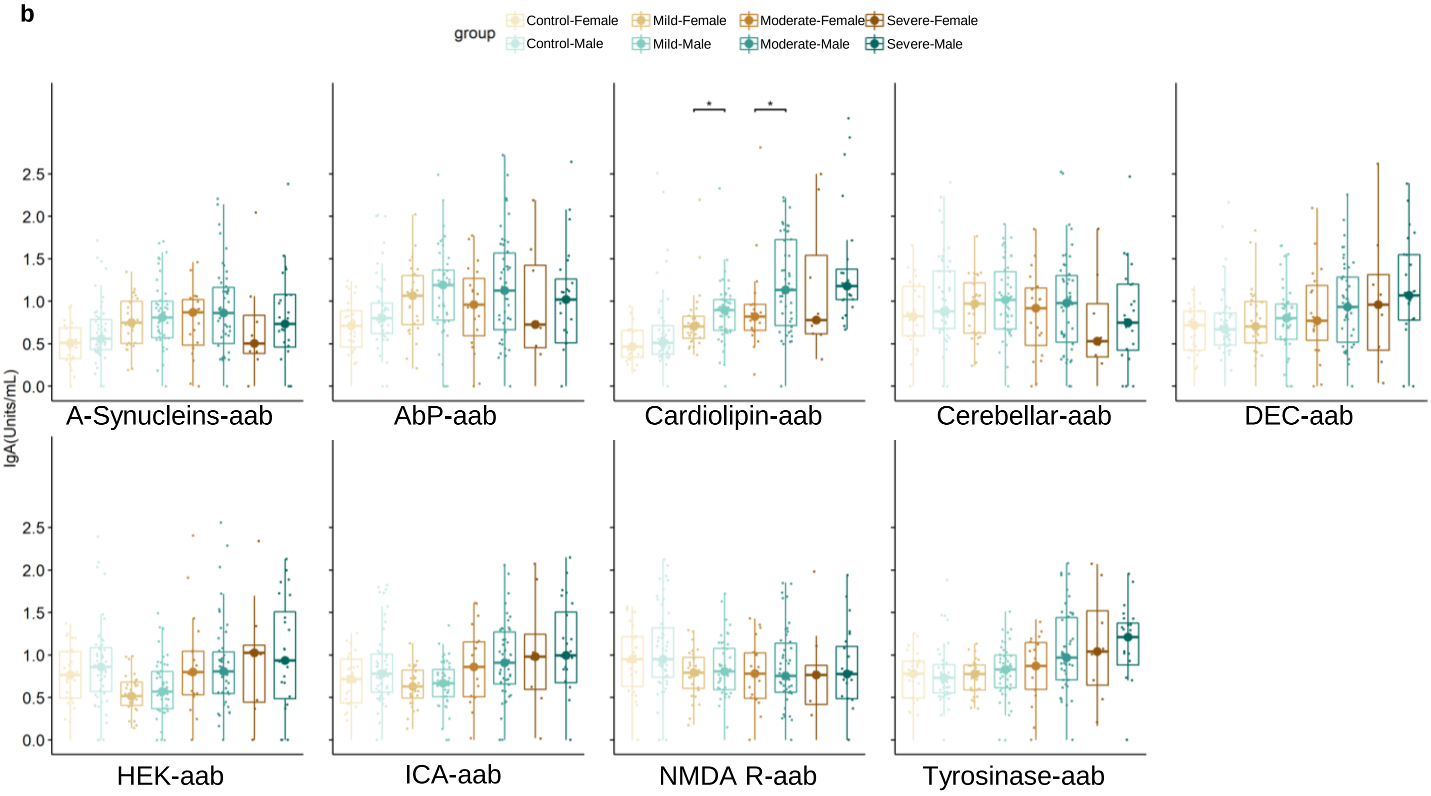
